# Sex-Stratified Genome-Wide Association Study of Multisite Chronic Pain in UK Biobank

**DOI:** 10.1101/2020.06.25.20140087

**Authors:** Keira JA Johnston, Joey Ward, Pradipta R Ray, Mark J Adams, Andrew M McIntosh, Blair H Smith, Rona J Strawbridge, Theodore J Price, Daniel J Smith, Barbara I Nicholl, Mark E.S Bailey

## Abstract

Chronic pain is highly prevalent worldwide and imparts significant socioeconomic and public health burden and is more prevalent in women than in men. Factors that influence susceptibility and mechanisms of chronic pain development, are not fully understood.

To investigate sex differences in chronic pain, we carried out a sex-stratified genome-wide association study of Multisite Chronic Pain (MCP), a derived chronic pain phenotype in UK Biobank. Genetic correlations between MCP in each sex and a range of psychiatric, autoimmune, and anthropometric phenotypes were examined. The relationship between female and male MCP, and chronic widespread pain was investigated using polygenic risk scoring. Expression of sex-specific MCP-associated loci in a range of tissues was examined using GTEx, and separately in neural and non-neural human tissues with assessment for dorsal-root ganglion (DRG) enrichment. For genes enriched for neural-tissue expression the full GTEx database was queried for sex-differential gene expression in CNS regions, and for high expression in sex-specific tissues. Expression in neural mouse tissue was also examined for orthologs of these genes.

A total of 123 SNPs at five independent loci were significantly associated with MCP in men. In women, a total of 286 genome-wide significant SNPs at ten independent loci were discovered. Meta-analysis of sex-stratified GWAS output found 87 independent SNPs to be significantly associated with MCP. We found sex-specific MCP-associated genes, with 31 genes and 37 genes associated with female and male MCP respectively and one gene associated with MCP in both sexes (*DCC*).

We found evidence for sex-specific pleiotropy and risk for MCP was found to be associated with chronic widespread pain in a sex-differential manner. Male and female MCP were highly genetically correlated, but at r_g_ significantly less than 1. All 37 male MCP-associated genes and all but one of 31 female MCP-associated genes were found to be expressed in the DRG, and many showed elevated expression in sex-specific tissues.

Overall, findings indicate sex differences in chronic pain at the SNP, gene and transcriptomic level, and highlight possible sex-specific pleiotropy for MCP. Results support the proposition of a strong nervous-system component to chronic pain in both sexes, emphasise the importance of the DRG, and indicate specific loci which may play a specialised role in nociception.

## Introduction

Chronic pain is widely defined as pain persisting beyond 3 months (1,2), and can be a primary disorder (3) or secondarily associated with injury, surgery or a range of medical conditions. Chronic pain is highly prevalent worldwide (4–9) and imparts a significant socioeconomic and public health burden (10). Factors influencing susceptibility to chronic pain, and the mechanisms underlying its development and maintenance, are not fully understood.

Several aspects of chronic pain including Chronic Pain Grade (11), severe chronic pain and low back pain have been studied from a genetic perspective and found to be complex traits. Heritability estimates vary from ∼30-46% in twin, pedigree and factor analysis studies (12–15), while single nucleotide polymorphism (SNP) heritability has been estimated from genome-wide association studies (GWAS) to be ∼7-10% (16,17).

It is increasingly recognised that sex differences in many complex human traits are biologically important, with genetic architecture for many traits being to some extent sex-specific (18), and a ‘sex-aware’ approach to genetic analysis has been widely advocated (19). Sex as a biological variable has wide ranging effects on the genome and on resultant phenotypes. These effects can be mediated via sex-differentiated gene expression (20,21), sex differences in methylation (22–26) and expression quantitative trait locus (eQTL) effects (27,28), or differing levels and actions of hormones (29,30). Sex can also influence traits through environmental factors strongly correlated with sex (23,24), and sex-specific pleiotropy (25,31). Chronic pain exhibits sex-related prevalence differences, and is more common in women than in men (32–34). There are also potential sex differences in the impact of pain on functioning in daily life, and in the success of specific coping strategies (35). In addition to differences in prevalence between the sexes, sex differences in underlying pain mechanisms and their modulation by immune cells have been recently reported (36,37), and immune responses in general can differ by sex (38).

Multisite Chronic Pain (MCP) is a derived chronic pain phenotype, defined as the sum of the number of sites of chronic pain on the body, on a scale from 0-7 (17). We have previously shown in UK Biobank (17) that genetic predisposition to MCP (as captured by a polygenic risk score; PRS) was associated with Chronic Widespread Pain (CWP), a separate but related chronic pain phenotype, in women but not men (17). Additional unpublished findings using the Generation Scotland study (39) demonstrated that the PRS-MCP was associated with both chronic pain grade and an MCP-like phenotype in both men and women, but that the magnitude of effect was roughly twice as great in women as it was in men. There was also a significant PRS-by-sex interaction. These findings suggest that sets of variants contributing to chronic pain in males and females may act differently, or have different genetic effect sizes, in the two sexes. Here we report on an exploration of this preliminary evidence for the existence of sex-specific loci associated with MCP using a sex-stratified GWAS analysis approach in UK Biobank, and identify several sex-specific MCP loci. A meta-analysis of the female- and male-specific GWASs also revealed novel MCP loci not identified in the original MCP GWAS. We have also investigated possible functional effects associated with sex-specific MCP-associated genes as revealed by gene expression data in multiple relevant tissues in both human and mouse.

## Methods

### Chronic Pain Phenotyping

UK Biobank participants were asked via a touchscreen questionnaire about “pain type(s) experienced in the last month” (field ID 6159), with possible answers: ‘None of the above’; ‘Prefer not to answer’; pain at seven different body sites (head, face, neck/shoulder, back, stomach/abdomen, hip, knee); or ‘all over the body’. The seven individual body-site pain options were not mutually exclusive, but those who chose ‘all over the body’ could not also select from the seven individual body sites. Where patients reported recent pain at one or more body sites, or all over the body, they were additionally asked (category ID 100048) whether this pain had lasted for 3 months or longer.

Chronic Widespread Pain (CWP) was defined as reported (40), and included only those participants who answered that they had pain ‘all over the body’ that was longer than 3 months in duration in the touchscreen questionnaire. These individuals were excluded from analyses of other chronic pain phenotypes as there is some evidence that this phenotype can be substantially different from more localised chronic pain (41). Multisite Chronic Pain (MCP) was a quasi-quantitative variable defined as previously reported (17); briefly, this variable captures the number of body sites at which chronic pain (at least 3 months duration) was recorded (excluding those with CWP): phenotypic values therefore ranged from 0 to 7. 10,000 randomly selected individuals reporting no chronic pain were excluded from the GWAS to use as controls in subsequent polygenic risk score (PRS) analyses.

### Genetic Quality Control

For the GWAS analyses, SNPs with an imputation quality score of less than 0.3, Minor Allele Frequency (MAF) < 0.01 and/or Hardy-Weinberg equilibrium (HWE) test p < 10^−6^ were excluded. Participants whose self-reported sex did not match their genetically determined sex, those who had putative sex-chromosome aneuploidy, those considered outliers in UK Biobank QC in terms of missingness or heterozygosity (42), and those who were not of self-reported white British ancestry were excluded from analyses. A summary of participant MCP phenotypic information for those included in each GWAS is shown in Table 1.

**Table 1:** number of participants per MCP phenotype level group included in each GWAS (male or female sex-stratified analysis).

| **MCP** | **N female** | **% female** | **N male** | **% male** |
| --- | --- | --- | --- | --- |
| 0 | 113148 | 54.11 | 105474 | 59.07 |
| 1 | 49984 | 23.91 | 42734 | 23.93 |
| 2 | 26000 | 12.43 | 18612 | 10.42 |
| 3 | 12376 | 5.92 | 7771 | 4.35 |
| 4 | 5319 | 2.54 | 2970 | 1.66 |
| 5 | 1723 | 0.82 | 780 | 0.44 |
| 6 | 471 | 0.23 | 181 | 0.10 |
| 7 | 72 | 0.03 | 34 | 0.02 |
| total n in each GWAS | 209093 | NA | 178556 | NA |

### BOLT-LMM GWAS

Sex-stratified GWASs of MCP, modelled as a quantitative trait, were carried out using BOLT-LMM (43), adjusting for age and chip (genotyping array), under the infinitesimal model of genetic risk, as previously described for our unstratified MCP GWAS (17). The SNP-level GWAS summary statistics were then analysed using FUMA (44) to obtain genome-wide plots and carry out MAGMA (45) gene-set and gene-based test analyses and gene expression analysis using GTEx (46) for male-specific & female-specific MCP loci. Significant independent and significant independent lead SNPs were determined according to FUMA. Briefly, FUMA defines lead SNPs as the subset of independent significant SNPs (SNPs associated with the trait at p < 5 × 10^−8^ and in LD at r^2^ < 0.6) which are in LD at r^2^ < 0.1 (44). In addition, when these LD blocks of independent significant SNPs are in close proximity (< 250kbp), they can be merged into a single genomic locus (each genomic risk locus can therefore contain multiple lead and independent significant SNPs) (44).

#### Meta-Analysis of Male and Female GWAS Summary Statistics

Meta-analysis of the two sex-specific GWAS summary statistics datasets was carried out using METAL (47), deploying a fixed effects model and inverse variance weighting. A meta-analysis p-value of < 5 × 10^−8^ was selected as the significance threshold for association.

### Transcriptome Analysis of Sex-Specific Association Gene Lists

We further analysed the tissue and cell type of expression of male-specific and female-specific genes from the MAGMA gene-based testing of sex-stratified GWAS results. Specifically, we characterized the gene expression in mammalian nervous system tissues and cell types as a potential starting point for identifying the functions of these genes with respect to pain. We also characterised expression for the one gene found to be associated with both male and female MCP in MAGMA analyses (*DCC*).

The Ray *et al* (48) study contains gene expression relative abundances in TPMs (Transcripts per Million) for 12 adult human tissues (6 neural and 6 non-neural). Additionally, the study notes 3 metrics on a scale of 0 to 1 for each gene based on expression in these 12 tissues : normalized Shannon’s entropy as a measure of tissue specificity (0 for highly tissue specific and 1 for tissue agnostic gene expression in the quantified tissues), neural proportion score as a measure of enriched expression (possibly neuronal and/or glial) in the nervous system (0 for genes not expressed in the nervous system and 1 for genes expressed solely in neural tissues with respect to these 12 tissues), and DRG enrichment score for identifying specificity of gene expression in the DRG with respect to the other 11 profiled tissues (0 or no expression in the DRG or for tissue-agnostic gene expression, and 1 for DRG specific gene expression in the context of the other profiled tissues). The mathematical formulations for these scores are provided in detail in Ray *et al* study (48). The corresponding tables are presented in Supplementary Tables 5 (male) and 6 (female).

Genes with neural proportion scores > 0.5 (with overall more neural than non-neural tissue expression), were further characterized by the putative cell type(s) of gene expression in the mammalian nervous system (Figure 4). While human single cell resolution RNA-seq datasets are not publicly available for the peripheral nervous system, a comprehensive database of gene expression in the mouse nervous system exists : the www.mousebrain.org repository (49). While it is true that nervous system expression patterns may be different between human and mouse between nervous system subpopulations due to regulatory evolution or due to differences in nervous system cell types (50), it is unlikely that expression would change categories across the 4 broad nervous system cell type categories : neurons, glia, immune and vascular cells) between human and mouse, given overall conservation of tissue gene expression profiles across humans and mice (48). A posterior probability characterizing the probability of detecting reads from a particular gene in a cell type subpopulation using a Bayesian framework, called the trinarization score is used with details in (49). Figure 4 visualizes trinarization scores for peripheral nervous system cell types sensory neurons and glia, enteric neurons and glia, and sympathetic neurons. While peripheral nervous system vascular and immune cells have not been profiled so far in www.mousebrain.org, we depict CNS vascular immune cells and vascular cells as surrogate cell types for their PNS counterparts. Both CNS neurons and glia play a critical role in chronic pain, but due to high diversity of cell types were not ideal for summarizing expression patterns in each subpopulation of these categories succinctly in a single figure. Instead, summary rows for CNS neurons and glia expression for relevant genes are provided at the bottom of the figure. For further details of expression in these subtypes, www.mousebrain.org can be queried for mouse gene expression profiles.

Finally, for genes with neural proportion scores > 0.5, the GTEx database was queried for sex differential gene expression in profiled CNS regions, and for high expression in sex-specific tissue like testis and ovary, noted in the comments section of Supplementary Tables 5 and 6.

### Genetic Correlations

Genetic correlations with a range of neuropsychiatric disorders and traits were assessed by LD-score regression (LDSR) for the male and female MCP GWAS outputs separately. Summary statistics datasets employed were publicly available or available via LD Hub (51)(52), or were results from published and unpublished in-house GWASs. LDSR p-values for genetic correlation were FDR-corrected within each sex. LDSR was also used to assess trait polygenicity and to calculate a SNP-heritability estimate. Note that although some of the GWAS used in this analysis came from studies that included UK Biobank data, genetic correlations estimated using LDSR are not subject to bias caused by sample overlap (53).

### Polygenic Risk Score Analysis

Previous analyses showed that a polygenic risk score (PRS) for MCP was associated with the phenotype of chronic widespread pain (CWP) in women but not in men, and that this PRS was associated more strongly with chronic pain phenotypes in women than in men in an independent cohort (Johnson K.J.A. et al., unpublished). To further explore the relationship between MCP and CWP, we assessed how separate male and female PRSs for MCP were associated with CWP. Separate sex-specific PRSs for MCP, based on the sex-specific GWAS results reported here, were calculated for a ‘case’ group consisting of participants who reported pain all over the body that lasted for three months or longer (a proxy phenotype for Chronic Widespread Pain; CWP; N = 6, 813)), and for a ‘control’ group consisting of 10,000 randomly selected UKB controls, both of which had been excluded from the GWAS analyses (demographic data for this subsample are given in Table 2)

**Table 2:** Summary of sample sizes of participants used in each of the two PRS analyses (male or female).

|  | Female |  | Male |  | Combined Totals |  |
| --- | --- | --- | --- | --- | --- | --- |
|  | N | mean age (years) | N | mean age | N | mean age |
| Control | 5135 | 56.68 | 4865 | 56.96 | 10000 | 56.81 |
| CWP | 4328 | 57.00 | 2485 | 57.27 | 6813 | 57.10 |
| Total | 9463 | 56.83 | 7350 | 57.07 | 16813 | 56.93 |

SNPs associated with MCP at p < 0.01 in the original sex-specific GWAS were selected and LD-pruned (at a threshold of r^2^ < 0.1 within a 250kbp window using PLINK ‘--clump’ command). Sex-specific PRS-MCPs were calculated for each individual in the analysis as the sum of risk alleles at each SNP, weighted by effect size (beta value) in the GWAS (54). PRS values were standardised by z-score for the analysis. Association between standardised sex-specific PRS-MCP and CWP status were investigated separately in males and in females in the target case-control subsample using logistic regression, adjusted for chip (genotyping array), age and the first eight genetic principal components.

## Results

### GWAS of MCP in males and females separately

To detect sex-specific genetic influences on multisite chronic pain, GWASs were run separately for males and females in UK Biobank. In men, a total of 123 SNPs at five independent loci were associated with MCP at a genome-wide significance threshold of p < 5 × 10^−8^ (Table 3, Figure 1). In women, a total of 286 genome-wide significant SNPs at ten independent loci were discovered (Table 3, Figure 1). The loci were differentially associated with sex - none of the SNPs at these loci had p < 5 × 10^−8^ in the GWAS conducted in the opposite sex. However, a total of 257 SNPs were found to have suggestive levels of association with MCP (p < 5 × 10^−5^) in both men and women. Two SNPs had p < 5 × 10^−5^ in men and were genome-wide significant in women, and eight SNPs had p < 5 × 10^−5^ in women and were genome-wide significant in men. In addition, the genome-wide significant loci on chromosome 6 in each sex were separated by less than 1 Mbp.

**Table 3:**
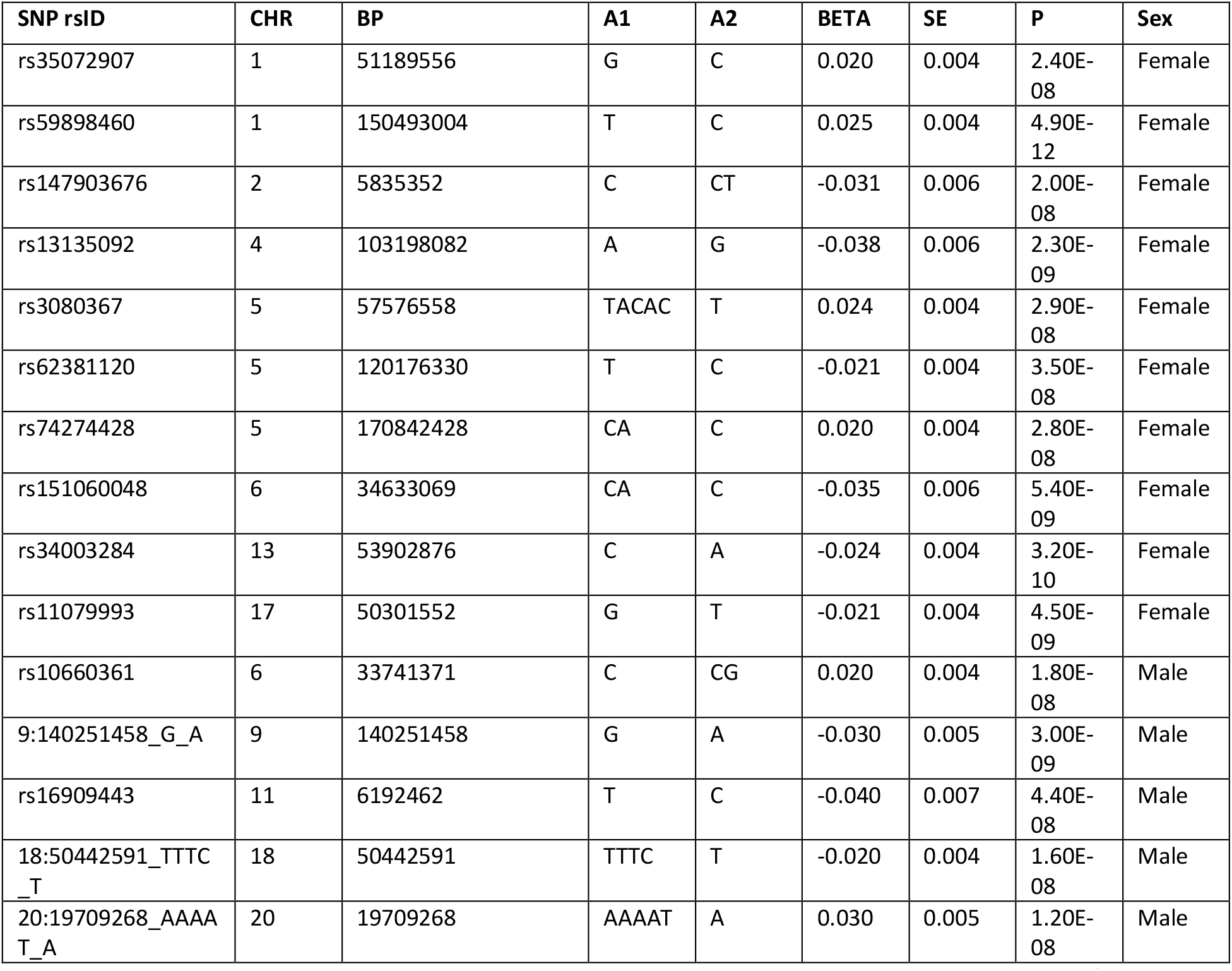
Genome-wide significant independent lead SNPs associated with MCP (p < 5 × 10^−8^) in male and female sex-stratified GWAS. Chr, chromosome; BP, chromosome coordinate; A1/A2, alleles 1 and 2, where A1 is the effect allele; Beta/SE, coefficient and its standard error for the effect allele.

**Fig 1.**
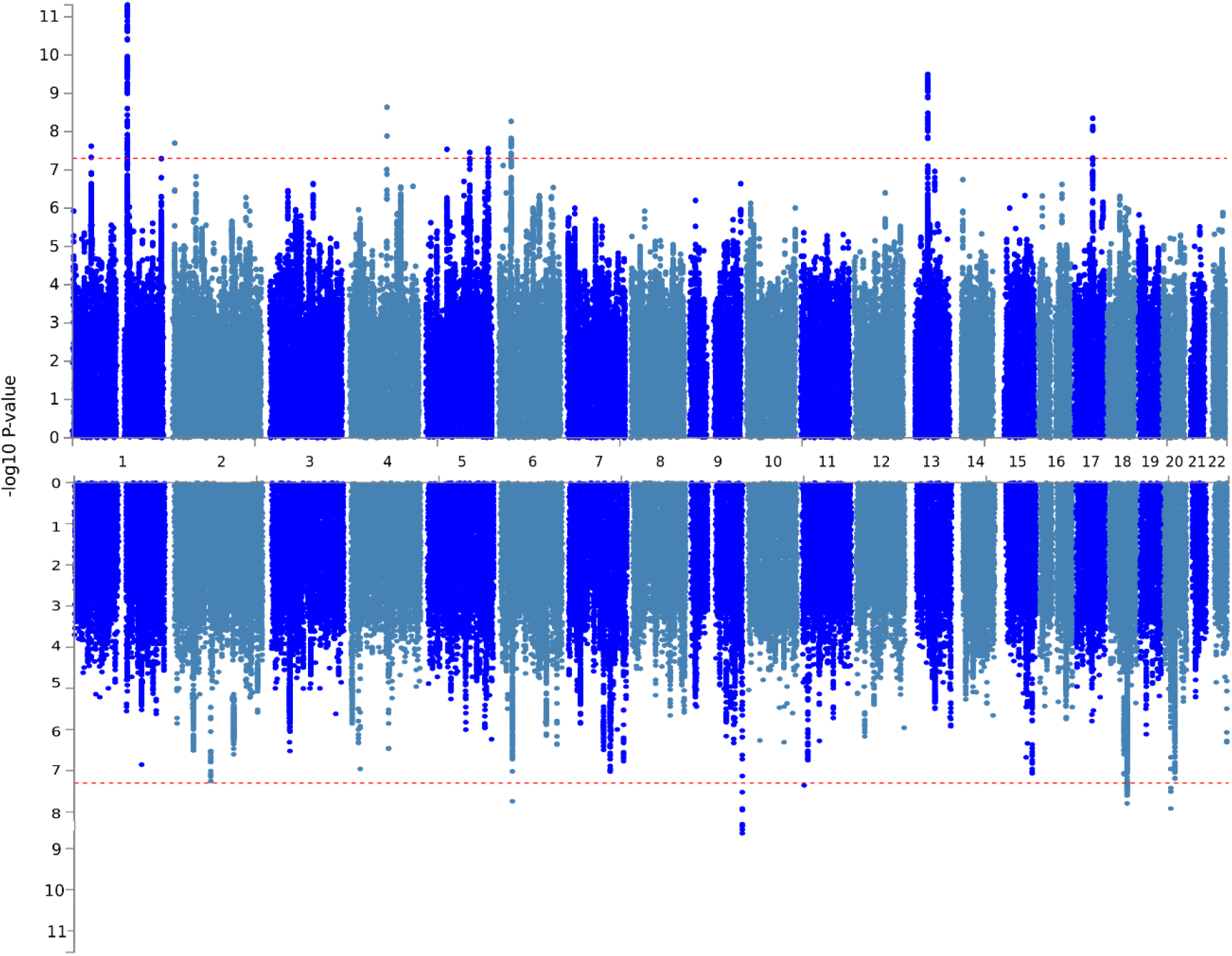
Manhattan plots for the stratified GWAS analyses. Upper panel: female, lower panel: male.

LDSR analysis demonstrated that inflation of test statistics in each of the two GWAS were due to polygenicity (Table 4; LDSR intercept). SNP heritability was moderate, estimated as 0.125 and 0.106 in females and males, respectively (Table 4). The genetic correlation between male and female MCP was high (r_g_=0.92, SE 0.03; p = 3.32 × 10^−213^), but significantly less than 100% (based on confidence intervals calculated as +/- 2 x SE).

**Table 4:** Trait genetic attributes from the male and female MCP GWASs. SNP-heritability, BOLT-LMM pseudo-heritability estimate,;lambda_GC1000_, lambda_GC_ value adjusted for sample size; LDSR_intercept (SE), LD-score regression intercept value and its standard error.

| Attribute | female | male |
| --- | --- | --- |
| SNP heritability | 0.125 | 0.106 |
| $\lambda_{GC1000}$ | 1.002 | 1.001 |
| LDSR_intercept (SE) | 1.03 (0.006) | 1.025 (0.005) |

### Meta-Analysis of Sex-Stratified MCP GWAS Outputs

In total, 49 lead SNPs across 46 genomic risk loci (Table 5) were found to be associated with MCP in meta-analysis of sex-stratified outputs. 87 independent SNPs were found to be associated with MCP at genome-wide significance in total.

**Table 5:**
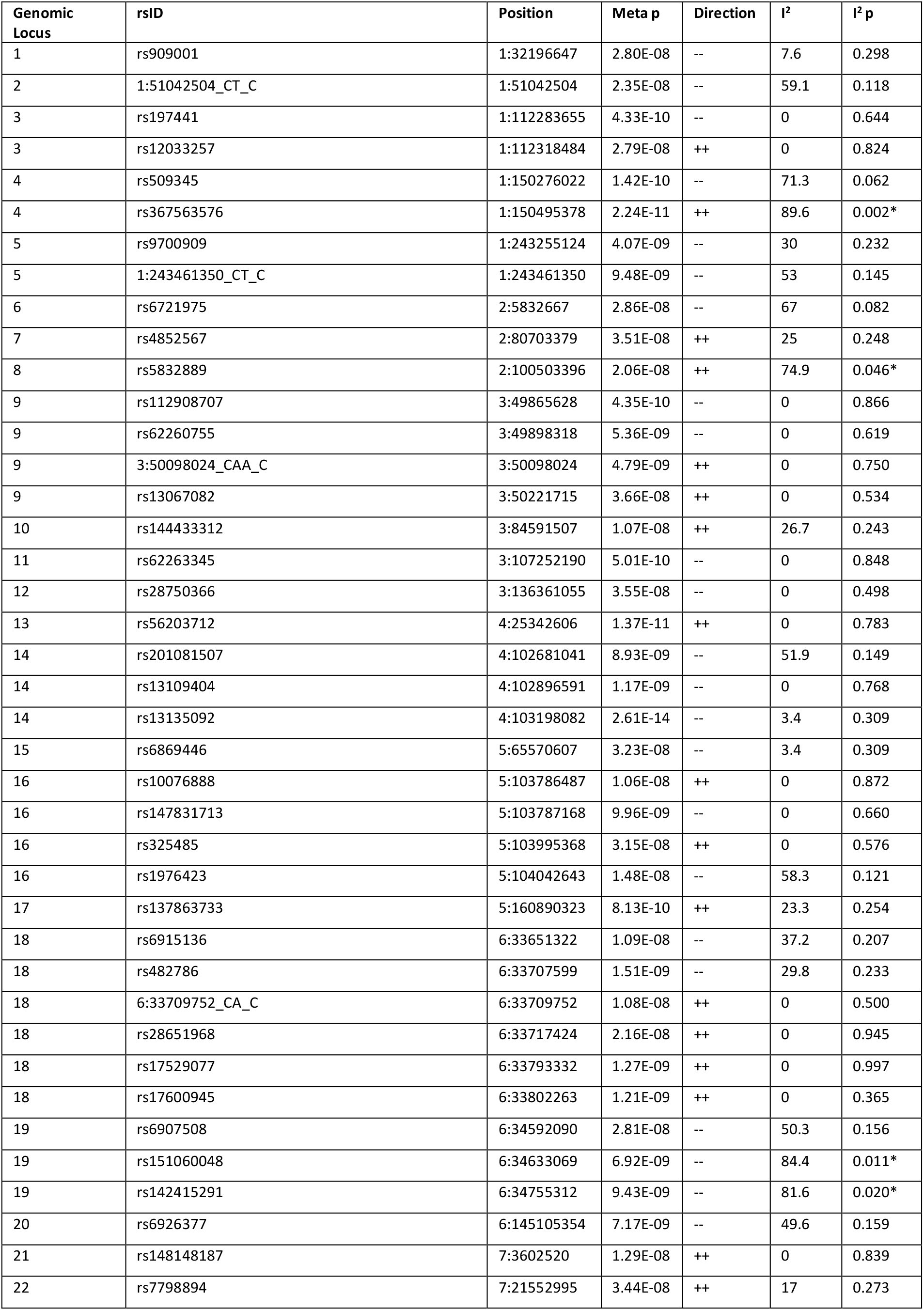
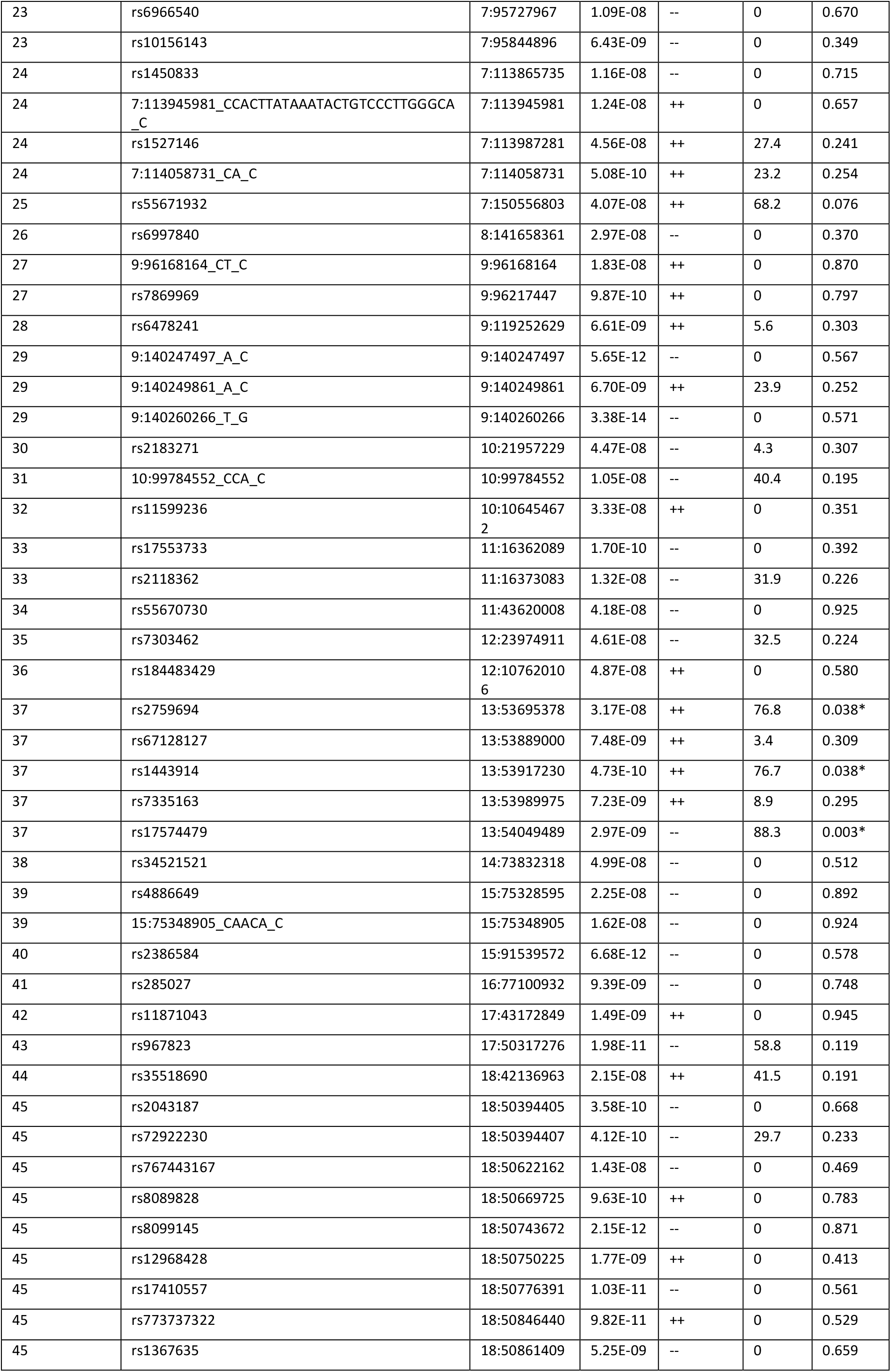
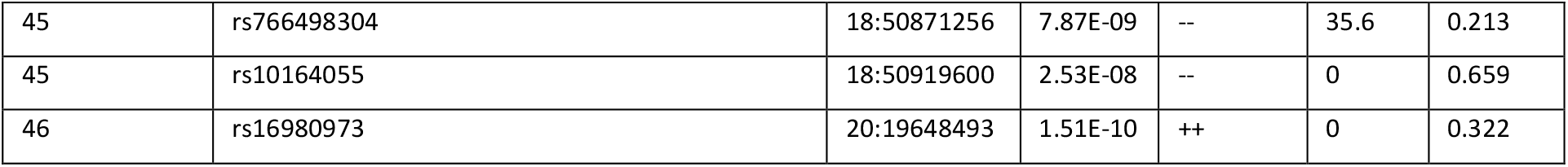
Independent, genome-wide significant SNPs from GWAS meta-analysis of sex-stratified MCP GWASs. Genomic Locus = genomic risk locus context of SNP, rsID = SNP rsID identifier, Position = genomic position (chromosome: base-pair start position), Meta p = p value for association for SNP from GWAS meta-analysis, Direction = direction of effect in female MCP and male MCP GWAS respectively (+ if association beta value for SNP > 0, - if < 0), I^2^ = I^2^ value (heterogeneity), I^2^ p = p value for heterogeneity measure. Significant heterogeneity I^2^ p values (I^2^ p < 0.05) are marked with *.

Each of the 87 independent significant SNPs showed consistent direction of effect between males and females (Table 5), but seven showed significant heterogeneity in effect size (I^2^ < 0.05).

### Genetic Correlations between sex-stratified MCP and other disorders and traits

LDSR analysis was carried out to determine genetic correlations between female and male MCP and a range of disorders and traits.

Results of LDSR analysis using summary statistics from the sex-stratified GWASs of MCP versus a range of potentially related disorders and traits. Genetic correlations are given as r_g_ values (and FDR-corrected p-values) sorted in order of numerically decreasing r_g_ for female MCP vs other traits. f_rg and m_rg = genetic correlation value for female and male MCP versus trait, respectively,, f_p_fdr and m_p_fdr = FDR-corrected p value for genetic correlation, source = source of trait GWAS data, PMID = pubmed ID of associated publication for GWAS of trait. Significant genetic correlations (FDR-corrected p value < 0.05) within each sex are highlighted orange, non-significant in blue.

MCP in both men and women was found to be significantly genetically correlated with a range of traits and disorders, including psychiatric and mood phenotypes such as anhedonia, mood instability, depressive symptoms, MDD, anxiety, suicidality and subjective wellbeing (Table 6). PTSD, schizophrenia, autism spectrum disorder, anorexia nervosa, PGC cross-disorder phenotype and primary biliary cirrhosis were found to be significantly correlated with MCP in one sex and not the other. Several phenotypes were not found to be genetically correlated with MCP in either sex (p_fdr_ >0.05), including inflammatory bowel diseases, Parkinson’s disease, bipolar disorder, rheumatoid arthritis and low relative amplitude.

**Table 6:**
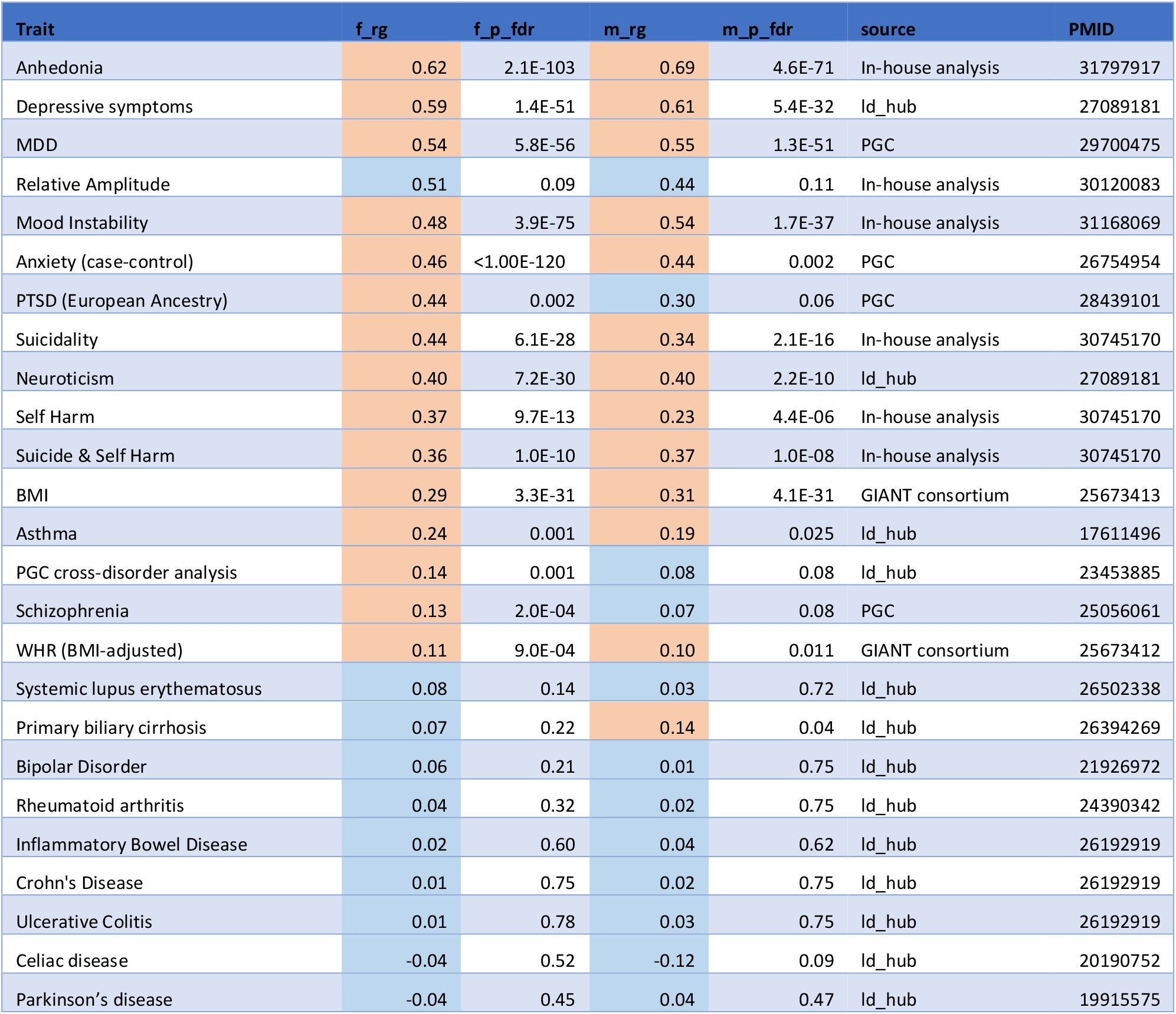
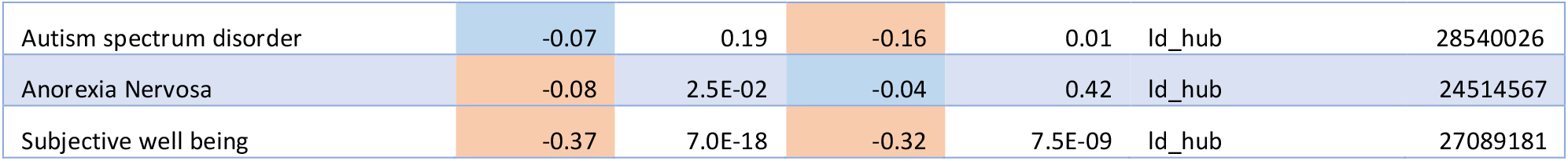
Genetic correlations between MCP and other disorders and traits

### Gene-level Analysis

Genes enriched for variants associated with MCP were identified using a gene-level association analysis (gene-based test) (45) approach implemented by MAGMA as part of the FUMA suite that tests 19, 012 separate genes. The results are summarised in Figure 2 and [Supplementary Table 1]. In females and males, 31 and 37 genes, respectively, were found to be significantly (Bonferroni-adjusted significance criterion: p < 2.63 × 10^−6^) associated with MCP. The only gene found to be significantly associated with both male and female MCP was *DCC*. 24 out of the 31 genes significantly associated with MCP in females in the sex-stratified analyses, and 31 out of the 37 genes significantly associated in males, were also significantly associated in our previous non-stratified analysis (17). Six genes were significantly associated with MCP only in females (*NCAN, SPATS2L, TBC1D9, CAMK1D, SOX11, GON4L*), while 4 genes were associated only in males (*CENPW, MTCH2, NICN1, DNAJA4*). Twenty-four genes were identified as significantly associated with MCP only in the meta-analysis of the sex-stratified GWAS outputs (Figure 2 and Supplementary Table 2).

**Figure 2:**
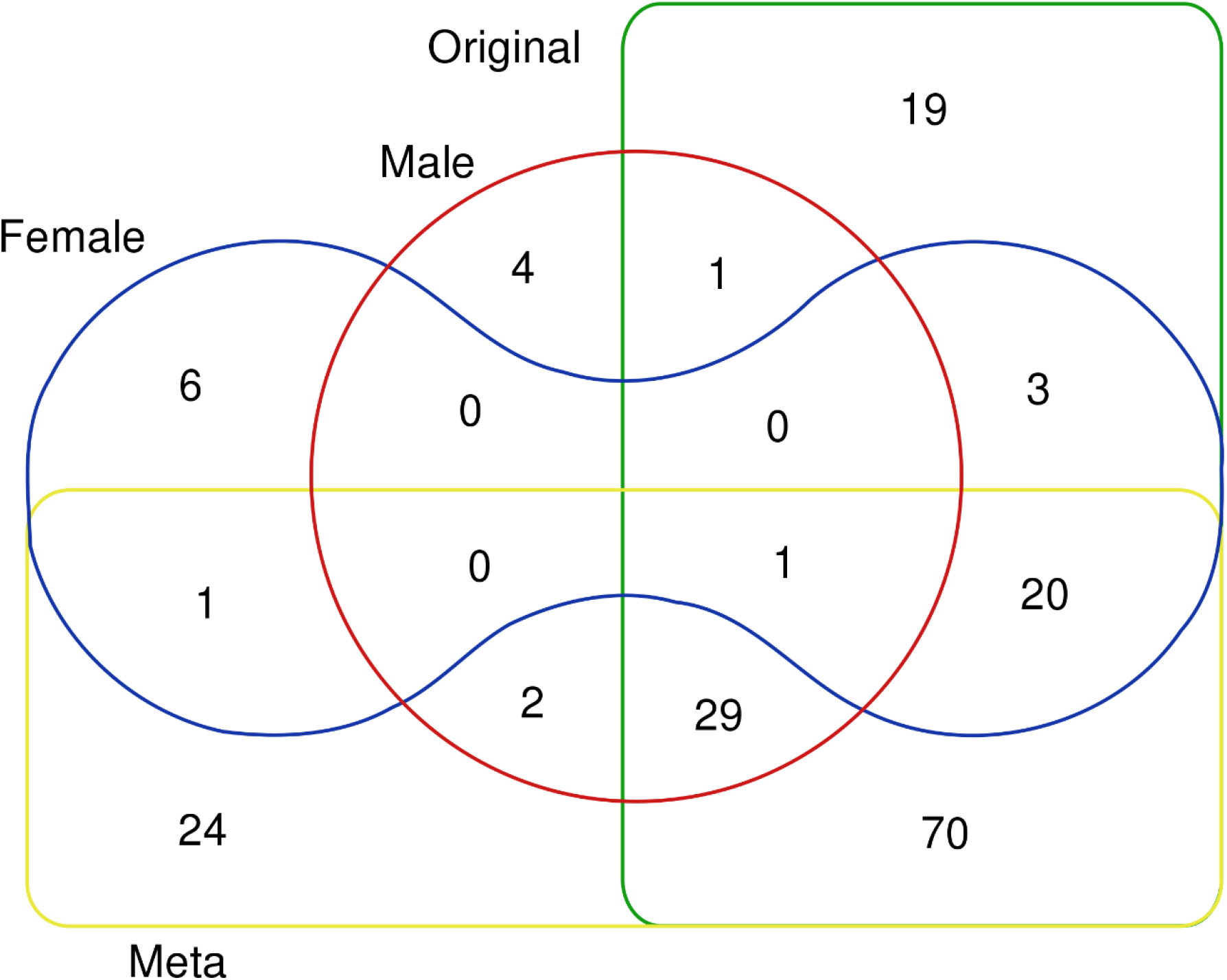
Venn diagram of number of genes found significantly associated in MAGMA gene-based test analyses. Sectors: Original = non-sex-stratified MCP GWAS, Meta = meta-analysed female and male GWAS output, Female = female MCP GWAS, Male = male MCP GWAS.

### Gene Expression Analysis

Association of GWAS results with tissue expression data was analysed using FUMA, which implements an MAGMA gene-property analyses (55) using GTEx (46) gene expression datasets to determine association between trait-associated genes and expression in a range of bodily tissues. MCP-associated genes in females were found to be enriched for expression the brain, particularly the cerebellum, cortex and frontal cortex (Fig 3). There was no tissue enrichment for expression of MCP genes found in males (Fig 3).

**Figure 3:**
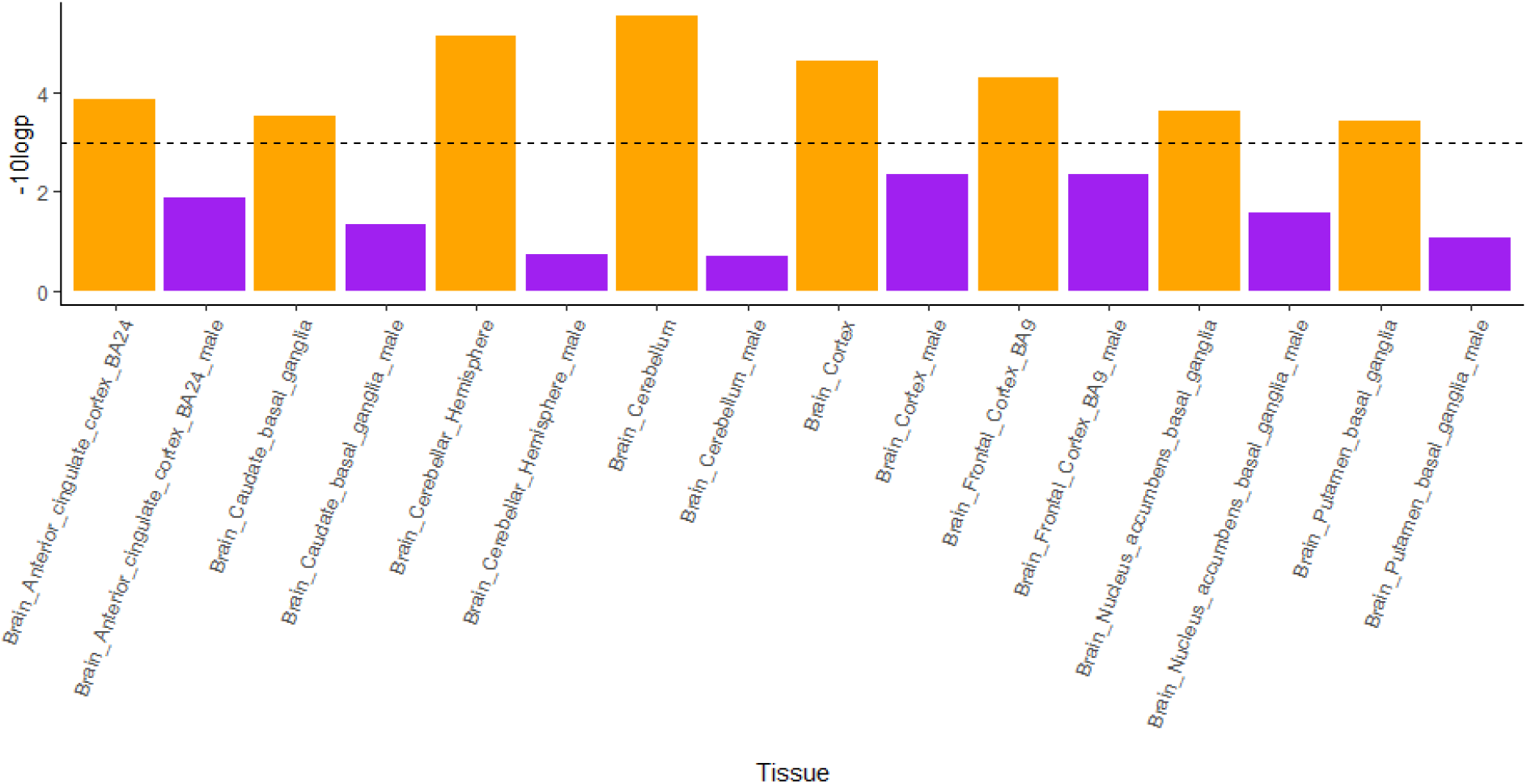
GTEx expression analysis results. Significant (−10logp > 3) results in female analyses and their corresponding tissue results in male analyses are shown for brevity, female in orange, male in purple. No significant enrichment for expression in any tissue was found for any MCP-associated gene in males. Dashed line = significance threshold. *Full GTEx results for all 53 tissues (8 of which shown here) can be found in [Supplementary Figures 1a & 1b].

For genes identified in the MAGMA gene-based testing analyses of sex-stratified GWAS outputs, we carried out further analyses of gene expression at tissue and cell-type level by querying existing transcriptomic datasets, focusing on neural tissues and tissues specific to each sex. For most tissues GTEx data were used, but because GTEx does not contain data for dorsal root ganglion (DRG) neurons, which are key for the generation of the nociceptive signals that initiate pain in chronic pain patients, we assessed expression in this tissue using other comparable datasets (48). We also used single cell sequencing datasets to estimate whether genes of interest are likely to be expressed in neurons in the peripheral or central nervous systems (CNS). Most of the 37 male-specific MCP-associated genes were observed to be expressed in the nervous system (see figure 4; full results are given in Supplementary Table 5). Two of the 37 genes, *IP6K3* and *FAM129A*, had low neural proportion scores, suggesting that they are non-neuronal and non-glial. All 37 genes, however, showed DRG expression, although expression level was very low for *DCC* and *IP6K3*, which were which were increased in expression in the CNS. One gene, *AMIGO3*, was found to be enriched solely in the DRG; its orthologue has also been found to be primarily expressed only in mouse DRG neurons in the www.mousebrain.org dataset (Figure 4).

**Figure 4:**
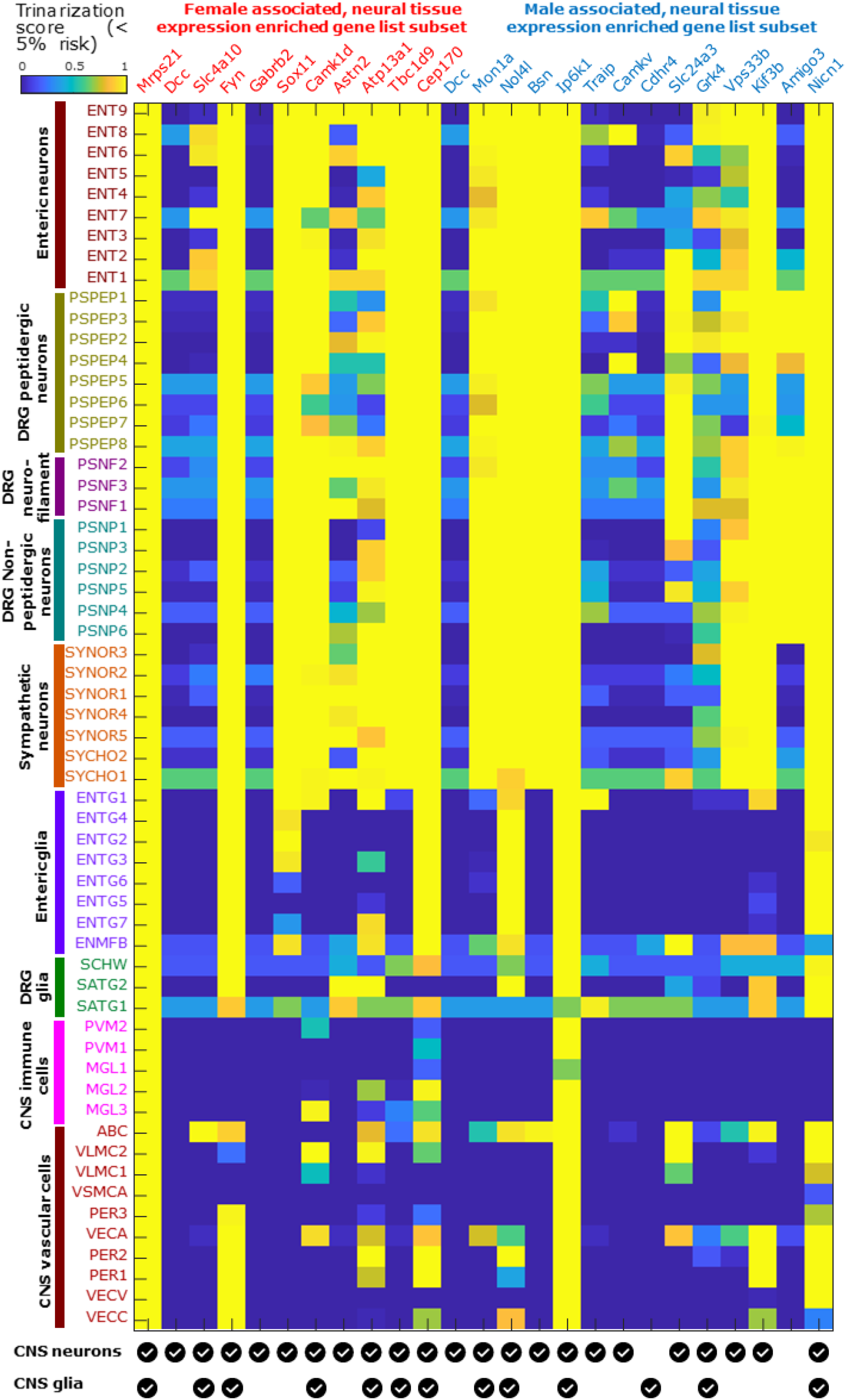
Mouse cell-type-specific expression patterns for sex-specific MCP-associated genes.4: The mousebrain.org database was used to identify cell types of expression in the mouse nervous system (brain neurons and glia have been collapsed to two lines at the base of the plot for clarity) for orthologues of genes identified in this study. Trinarization score (posterior probability of detection in a particular cell type) was used, rather than expression levels. The analysis was carried out for 25 genes selected from the male-specific and female-specific MCP-associated sets as being enriched for neural tissue expression (neural proportion score > 0.5, Supplementary tables 5 and 6). While most of these show pan-neuronal expression, a few are expressed in mouse glial subpopulations (*Cdhr4*), primarily in CNS neurons (*Gabrb2*), primarily in DRG neurons (*Amigo3*), or in limited subsets of neurons (*Dcc, Slc4a10, Camkv*).

Of the 30 female-specific MCP-associated genes, all showed high neural expression except *CPS1*, which was not expressed in neural tissue at all. This gene and several others that were expressed in neural tissues but with low (< 0.5) neural proportion scores (e.g. *SEMA3F, MST1, MST1R, SDK1, ECM1*; supplementary tables 5 and 6) were found to be involved in immune function. Among the genes with high (> 0.5) neural proportion scores, mouse orthologues of many are pan-neuronal in expression based on the mousebrain.org dataset (figure 4), with a few genes having ubiquitous but neural-enriched expression (*Mrps21, Ip6k1*), solely glial expression (*Cdhr4*), or being expressed in limited neuronal subpopulations including CNS, sensory and enteric neuronal subtypes (*Dcc, Camkv, Slc4a10* respectively). Several of the neuronally expressed genes are known to be involved in axon pathfinding and neurite outgrowth. Many of these genes have elevated expression levels in sex-specific tissues like testis and ovary, and / or are somewhat differentially expressed (10% or more change in median TPMs across sexes) between male and female cohorts in brain sub-regions in the GTEx database (supplementary tables 5 and 6), suggesting they may be androgen- or estrogen-regulated. All 30 genes except *GABRB2* (whose mouse ortholog is primarily expressed in CNS neurons, Figure 4) did show expression in DRG (supplementary table 6), though none of them showed enriched expression in the DRG compared to the CNS.

### Polygenic Risk Score Analysis

Polygenic risk scores were used to assess whether MCP and chronic widespread pain (CWP), a related but distinct chronic pain phenotype, are likely to be related biologically. For both men and women, the sex-specific PRS-MCP was significantly associated with CWP (p << 0.05; O.R. = 1.0034 (female) and 1.0026 (male); [Supplementary Tables 3 & 4], indicating that genetic risk for MCP is significantly associated with having chronic widespread pain, and that the degree of association is bigger in women than in men.

## Discussion

### Comparing Male and Female Multisite Chronic Pain

Prevalence, coping strategies, and, potentially, mechanisms of development and maintenance of chronic pain vary between the sexes. To explore underlying genetic differences that may contribute to these sex differences in chronic pain, we carried out a large-scale sex-stratified GWAS of a quantitative chronic pain phenotype, MCP. We found both male and female MCP to be moderately heritable. Although the estimated female SNP heritability was higher than that in males (12.5% versus 10.6% respectively), this difference was not significant.

#### MCP-Associated SNP Loci

Twice as many genomic risk loci were identified in GWAS analyses in females as in males (10 versus 5 respectively), with no risk loci shared between the sexes. This may be due to lower sample size in the male MCP GWAS, both in comparison to previous non-stratified analyses and to sample size in female MCP GWAS analysis (roughly 30,000 more participants are included in the female MCP GWAS than in the male GWAS). Loci were found across the genome (Fig 1., Table 3) in both males and females, with genomic location varying by sex. Significant loci were found in both men and women on chromosome 6, 0.89 Mbp apart (Table 3) but with different directions of effect (trait-increasing in men, trait-decreasing in women).

Additional trait-associated SNPs were discovered when the sex-stratified GWAS outputs were meta-analysed, probably as a consequence of increased power. Twenty-four loci that had not reached genome-wide significance in our previous sex-combined analysis were found to be associated with MCP after meta-analysis. The fact that these loci were not identified in the non-stratified GWAS could be due to effect heterogeneity, in terms either of direction or magnitude, between the sexes reducing the overall signal at these loci.

Previous studies have highlighted sex-specific loci and loci with heterogeneous effects between the sexes in a range of disorders and traits, such as ASD, anthropometric traits and asthma (31,56–58). Four of the loci found to be associated with MCP in our meta-analysis of sex-stratified GWAS outputs, including 7 SNPs in total, showed significant heterogeneity of effect size between the sexes, and these loci may contribute to sex differences in chronic pain. Previous studies have highlighted sex-heterogeneous or sex-specific loci in a range of disorders and traits, such as ASD, anthropometric traits and asthma (31,56–58). A total of seven of the 87 significant, independent SNPs found to be associated with MCP in meta-analysis of sex-stratified GWAS outputs showed significant heterogeneity between the sexes, across four different genomic risk loci, and these loci may contribute to a genetic component in sex differences in chronic pain.

#### Genes of Interest

##### Genes Associated with MCP in Males

Genes found to be associated with MCP in males in the stratified GWAS included *CENPW, MTCH2, NICN1, AMIGO3, DNAJA4, CTBP2* and *NOP14*, with the latter two also being significant in the meta-analysis. *CENPW* encodes centromere protein W, involved in kinetochore assembly and function, and associated with diseases such as type 1 diabetes (59–61). *MTCH2* (mitochondrial carrier 2) encodes a member of the SLC25 family, a family of transporters localised to the inner membrane of mitochondria and involved in a wide range of cell metabolism functions (62). SNPs in this locus have previously been associated with obesity (63–65), and this gene may be involved in regulation of development of adipocytes. *NICN1* (nicolin 1) encodes a nuclear protein of unknown function expressed in a variety of tissues (66). *AMIGO3* (adhesion molecule with Ig-like domain 3), the only male-specific MCP-associated gene whose expression was enriched in DRG, is a member of a small family of cell-surface immunoglobulin domain- and leucine-rich repeat-containing adhesion molecules. Its function is not well understood, but it is expressed in a range of DRG neuronal subtypes and may play a specialized role in nociception or other sensory modalities. Interestingly, *AMIGO3* is located almost next to and within 40 kbp of *IP6K1*, which was also found to be associated in the male gene-level analysis (Suppl. Table 1). It may be that there is coordinate regulation of these two genes in tissues relevant to MCP, or that a number of separate functional variants are distributed across this genomic locus but in fact only influence expression of one of these two genes.

*DNAJA4* (DnaJ Heat Shock Protein Family (Hsp40) Member A4) encodes a heat shock protein (67) previously shown to be involved in melanoma metastasis and angiogenesis regulation, but is generally poorly characterised (68).*CENPW* is a protein coding gene (Centromere Protein W), involved in kinetochore assembly and function, and associated with diseases such as diabetes (Type 1) (59–61). *MTCH2* (Mitochondrial Carrier 2) encodes a member of the SLC25 family, a family of nuclear encoded transporters localised to the inner membrane of mitochondria and involved in a wide range of cell metabolism functions (62). SNPs in this locus have been previously associated with obesity (63–65), and this gene may be involved in regulation of development of adipocytes. The gene *NICN1* (Nicolin 1) encodes a nuclear protein of unknown function expressed in a variety of tissues (66). The function of *AMIGO3* is not well known, but this was the only gene that showed enriched expression in the DRG suggesting it may play a specialized role in nociception. *DNAJA4* (DnaJ Heat Shock Protein Family (Hsp40) Member A4) encodes a heat shock protein (67), previously shown to be involved in melanoma metastasis and angiogenesis regulation but is generally poorly characterised (68).

##### Genes Associated with MCP in Females

Genes found to be associated with MCP in females included *NCAN, SPATS2L, TBC1D9, CAMK1D, SOX11, GON4L*, and *DAGLB*, the last of which was also significantly associated in the meta-analysis. *NCAN* (neurocan) encodes a chondroitin sulfate proteoglycan (69) potentially involved in the modulation of cell adhesion and migration, and previously linked to bipolar disorder in GWAS and mouse model studies (70,71). *SPATS2L* (spermatogenesis associated serine rich 2 like) encodes a protein that may be involved in ribosome biogenesis and translational control as a response to oxidative cellular stress (72). *TBC1D9* (TBC1 domain family member 9) encodes a potential GTPase and was found to be overexpressed in mantle cell lymphoma (73). *TBC1D9* was also recently found to be involved in a Ca^2+^-dependent cellular response to infection (74). *CAMK1D* (calcium/calmodulin dependent protein kinase ID) encodes a member of the calcium/calmodulin-dependent protein kinase 1 family involved in granulocyte regulation, activating CREB-dependent gene transcription, the activation and differentiation of neutrophils, promotion of basal dendritic growth of hippocampal neurons, and apoptosis in erythroleukemia cells (75). *SOX11* (SRY-box transcription factor 11) encodes a member of the SOX (SRY-related HMG-box) family of transcription factors, with potential roles both in nervous system development and in neurogenesis during adulthood (76–80). *De novo* mutations in this gene have also been associated with Coffin-Siris syndrome (81). *GON4L* (Gon-4 like) encodes a protein involved in transcriptional repression (82,83). *DAGLB* (diacylglycerol lipase beta) encodes an enzyme that participates in the endocannabinoid synthesis pathway and is required for axonal growth during development and for retrograde synaptic signalling in mature synapses (84).*NCAN* (Neurocan) encodes a chondroitin sulfate proteoglycan (69) potentially involved in the modulation of cell adhesion and migration, and previously linked to bipolar disorder in GWAS and mouse model studies (70,71). *SPATS2L* (Spermatogenesis Associated Serine Rich 2 Like) encodes a protein which may be involved in ribosome biogenesis and translational control as response to oxidative cellular stress (72). *TBC1D9* (TBC1 Domain Family Member 9) encodes a protein which may function as a GTPase and was found to be overexpressed in mantle cell lymphoma (73). *TBC1D9* was also recently found to be involved in Ca2+-dependent cellular response to infection (74). *CAMK1D* (Calcium/Calmodulin Dependent Protein Kinase ID) encodes a member of the calcium/calmodulin-dependent protein kinase 1 family, a subfamily of the serine/threonine kinases, involved in granulocyte regulation, activating CREB-dependent gene transcription, the activation and differentiation of neutrophils, promotion of basal dendritic growth of hippocampal neurons, and apoptosis in erythroleukemia cells (75). *SOX11* (SRY-Box Transcription Factor 11) encodes a member of the SOX (SRY-related HMG-box) family of transcription factors which are involved in regulating embryonic development, including potentially regulating development of the nervous system and in neurogenesis in adulthood (76–80). Mutations (*de novo*) in this gene have also been associated with Coffin-Siris syndrome (81). *GON4L* (Gon-4 Like) encodes a protein involved in transcriptional repression (82,83). *DAGLB* (Diacylglycerol Lipase Beta) encodes a protein catalyst of endocannabinoid production, required for axonal growth during development and retrograde synaptic signalling in mature synapses (84).

Only one gene, *DCC* (DCC netrin 1 receptor; a.k.a. deleted in colorectal carcinoma), was associated with both male and female MCP. *DCC* encodes a receptor for the guidance cue netrin 1, and is important for development of the nervous system, particularly the dopaminergic system (85).Overall, genes associated with male and female MCP differed (with the only gene associated with both male and female MCP being *DCC*). *DCC* (Deleted in Colorectal Cancer a.k.a. DCC netrin 1 receptor) encodes *DCC*, the receptor for the guidance cue netrin 1, which important for development of the nervous system, particularly dopaminergic pathways (85). This receptor has also been linked to tumorigenesis in cancers other than colorectal (86), and mutations in the *DCC* gene have been found in those with congenital mirror movement disorder (87).

#### Gene Expression Differences in Male and Female MCP

Gene expression analyses in GTEx (which does not have DRG samples) carried out using FUMA indicated that expression of the female-specific MCP-associated genes was enriched primarily in brain tissue, and this pattern was also seen when meta-analysed sex-stratified GWAS outputs were analysed similarly (not shown). Almost all of these genes were also expressed in the human DRG suggesting that cells located within this structure may also play a significant role in initiation or maintenance of chronic pain phenotypes. No significant enrichment for specific tissues was seen in analyses of male MCP-associated genes using GTEx and the human DRG expression profiles. The lack of tissue-enrichment findings for the male-specific genes may, as with the lower number of MCP-associated loci, be due to reduced power resulting from lower sample size in the male GWAS. However, these patterns of tissue-level gene expression in GTEx may also indicate differing gene expression between the sexes, with more ubiquitous expression across all tissues for genes associated with MCP in males, while genes conferring risk in females may tend to have more tissue-specific expression patterns. These expression patterns may also be associated with the fact that the GTEx resource is enriched for male tissue samples (V8 release; 67.1% male) – sex-differential enrichment of certain genes may be conflated with tissue tissue-differential gene enrichment.

It is still notable that almost all the genes in male or female MCP-associated genes were found to be expressed in human CNS tissues and in DRG. A subset of these are enriched in human neural tissues and additionally are expressed in mouse neuronal subpopulations when examining single cell sequencing databases (Figure 4). All of these evidences, together, suggests putative central and peripheral neuronal roles for some of these genes, many of which have not been historically well studied in the field of chronic pain.

#### Genetic Correlations

For both males and females, MCP was genetically correlated with non-stratified MCP (17), at r_g_= 1. In contrast, the genetic correlation between male MCP and female MCP was found to be high but significantly less than 1 (r_g_= 0.92), with around 8% of common SNP-tagged trait variation therefore not shared between the two traits. However, it could be argued that although this figure suggests that a small subset of genetic variation linked to MCP is unique to each sex, this difference is not large enough to consider MCP in the two sexes as biologically distinct to a considerable extent -- traits correlated at lower r_g_are routinely used as proxies for one another in GWAS settings e.g. educational attainment as proxy for intelligence (r_g_∼ 70%) (88), or current age as a proxy for life span (r_g_∼40-70%) (89).

A range of complex trait phenotypes were selected for LDSR analysis with male and female MCP based on previous evidence for phenotypic correlation (17), with the addition of newly available trait data such as GWAS outputs on suicide and self-harm (90) and mood instability (91). Suicidality and self-harm are important comorbidities of chronic pain, an issue compounded by the common co-occurrence of mental health traits such as MDD with chronic pain, and of certain medication usage in chronic pain both contributing to increased risk for self-harm, suicidal ideation and suicide attempt (92–97). When GWAS outputs from each sex-stratified analyses were used to examine sex-specific genetic correlations between this range of traits, several key differences were observed: there were significant genetic correlations between certain psychiatric and mental health traits and female MCP but not male MCP, plus significant genetic correlations between certain autoimmune and neurodevelopmental disorders and male MCP but not female MCP (Table 5)

Significant genetic correlations of similar magnitude for both sexes were found between MCP and anhedonia (r_g_>0.6), mood instability (r_g_∼0.5), MDD (r_g_∼0.55), depressive symptoms (r_g_∼0.6), BMI and WHR adjusted for BMI (r_g_∼0.10-0.30), anxiety (r_g_∼0.45), asthma (r_g_∼0.20), neuroticism (r_g_=0.4), suicidality (r_g_∼0.4), subjective wellbeing (r_g_∼ -0.35) and self-harm (r_g_∼0.20-0.4). No significant genetic correlations were found between MCP in either sex and some neuropsychiatric conditions (BD), some physiological/circadian traits (relative amplitude) and several inflammation/immune system-related conditions (rheumatoid arthritis, SLE, Parkinson’s disease, celiac disease, inflammatory bowel disease including Crohn’s disease and ulcerative colitis).

Significant genetic correlation was seen between female MCP and schizophrenia (r_g_=0.13), PGC cross-disorder phenotype (r_g_=0.14), PTSD (r_g_=0.44) and anorexia nervosa (r_g_=-0.08), whilst no significant genetic correlation was observed between these traits and MCP in males. In contrast, significant genetic correlation was observed between MCP in males (but not females) and autism spectrum disorder (r_g_=-0.16), and male MCP and primary biliary cholangitis (r_g_=0.14).

This may indicate sex-specific differences in pleiotropy, with potentially different underlying genetic variation associated with both these traits and MCP in a sex-dependent manner. In particular, pleiotropic variants may contribute jointly to a greater degree to variation in particular psychiatric disorders (schizophrenia, PTSD) and female MCP, but not male MCP.

However, observed differences in pleiotropy may be due to differences in sample size between male and female MCP GWASs (with lower sample size for men compared to women). Differences may also be a result of either men or women being over-represented in certain GWASs used for LDSR cross-trait analyses. For example, in the GWAS meta-analysis of autism spectrum disorder (PMID 28540026) contributing cohorts had M:F ratios from 1.2:1 to as high as 8.6:1, and the anorexia nervosa GWAS (PMID 24514567) contained only female cases.

In contrast, this does not appear to be the case for PTSD, which was significantly correlated with MCP in females and not males. M:F ratios for the subsamples of cohorts included in the PTSD meta-analysis were not directly provided (PMID 28439101) and are not always discernible from the original cohort publications or cohort-related publications. For example, although White/ White-European ancestry participants were included in PTSD GWAS meta-analyses which were then used for LDSR, demographic information including sex ratio is given on the whole mixed-ancestry cohort in the original publication. In other instances, a PTSD-endorsing subsample of a wider cohort is used in GWAS meta-analysis but sex ratio is only given for other substance use case/ control participant groups. Despite this, it was possible to roughly estimate M:F ratios in cohorts contributing to the PTSD GWAS used in LDSR analyses here. Of the seven cohorts, some appear to be all-female (Nurses Health Study II (98)), predominantly female (Grady Trauma Project (99)), or roughly 1:1 (Family Study of Cocaine Dependence (100), Collaborative Study of Nicotine Dependence (101)). The remaining three cohorts used in the European-ancestry PTSD GWAS meta-analysis were all-male (Marine Resiliency Study (102)) or predominantly male (Ohio National Guard (99), Mid-Atlantic Mental Illness Research Education and Clinical Center PTSD Study (103), VA Boston-National Center for PTSD Study (104)). This suggests the difference in genetic correlation with MCP (significant in females and not males) is not driven by an overrepresentation of females in the PTSD GWAS used in LDSR analyses.

Similarly, although individual cohorts contributing to the schizophrenia GWAS meta-analysis used here (PMDI 25056061) ranged in % male participants from 26.8-91.8%, mean percentage of male participants across all cohorts in the European ancestry subset was 58%, again suggesting differences in genetic correlation with MCP between the sexes are not driven by overrepresentation of females in the schizophrenia GWAS meta-analysis.

Overall, when the correlations are visualised as in Fig 5, a general similarity in correlations between MCP and other disorders is seen in men and women, even though some are non-significant in one sex and not the other. There is however stronger evidence for sex-specific pleiotropy in terms of the relationship between female MCP and PTSD, and between female MCP and schizophrenia.

**Figure 5:**
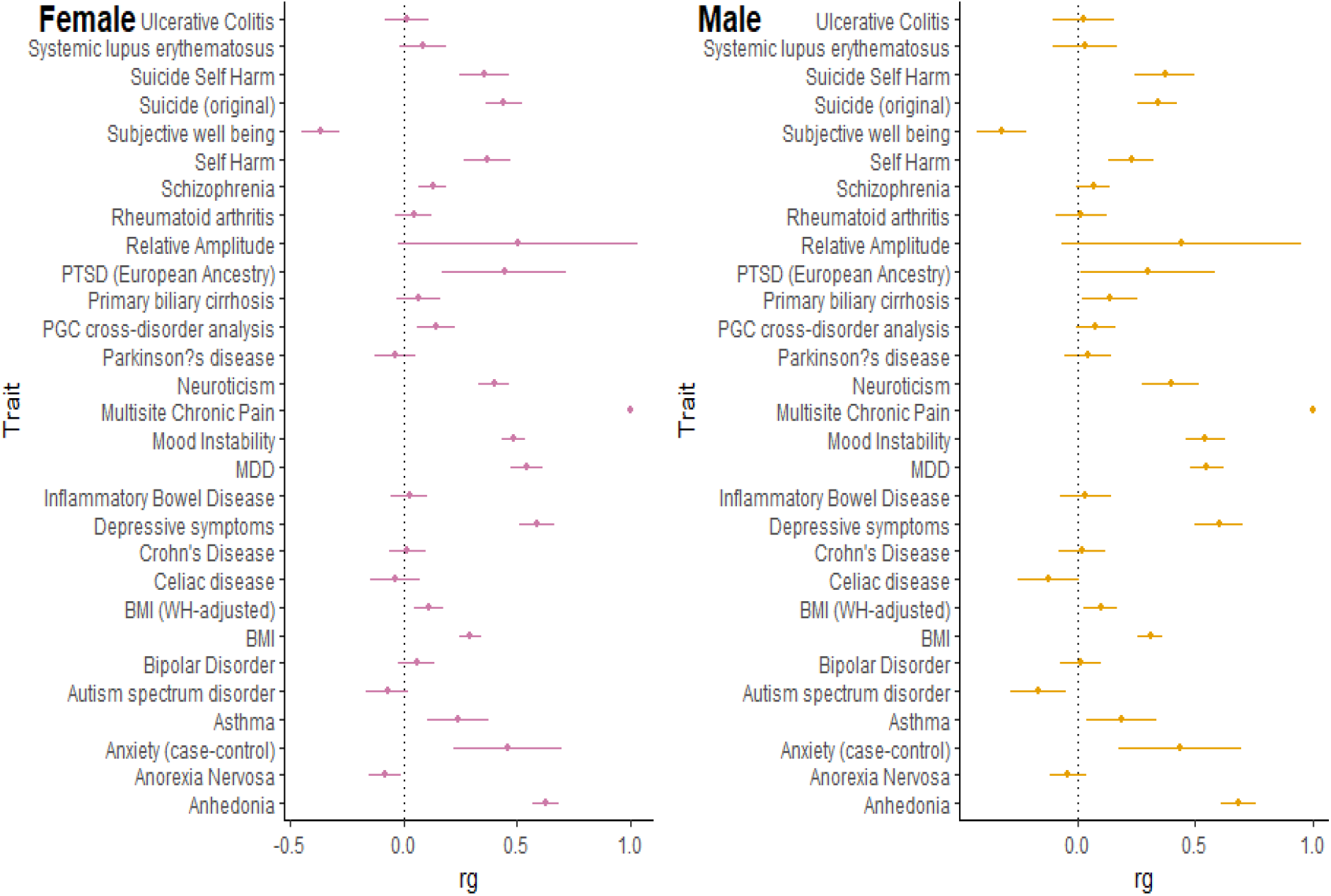
Genetic correlations between MCP and multiple traits in women and men (r_g_ values with 95% CI error bars).

#### Comparing the Relationship between Chronic Widespread Pain and MCP in Males and Females

Each sex-specific PRS was significantly associated with CWP in the corresponding sex, but the magnitude of association was much lower in comparison to the sex-combined PRS analysis reported previously (17), which may reflect the smaller sample sizes in the sex-stratified analyses. As previously found in sex combined analyses, results indicated a moderate degree of shared genetic basis for MCP and chronic widespread pain in both sexes, though potentially a slightly stronger shared genetic basis in females. It is possible that this difference is driven at least in part by the overrepresentation of females in CWP cases (M:F ratio 1:1.74).

#### Clinical Perspective on Findings and Potential Impact on Treatment

These results suggest that MCP shows genetic (and therefore biological) differences between men and women. If women with MCP are more likely to be at risk of PTSD, schizophrenia, and eating disorders, specific screening for these could be appropriate, with a view to instigating additional appropriate management or referrals. Except for PTSD, however, these sex-specific genetic correlations were relatively small in comparison with those that affected both sexes, particularly a range of mood disorders. It remains essential to assess the possibility of these in all patients with MCP, and to instigate appropriate management.

While there are clearly also many similarities, it is important that clinicians begin to recognize these differences in clinical assessment and awareness of risks. While more research is required, these differences may also eventually inform choice of medical treatment. For example, if more MCP in women is associated with immune function, drugs that influence this may have specific long-term effects (positive or negative) in women and not in men. Opioids are known to adversely affect immune function (105,106). These differences may also inform the search for new, or re-purposed drugs in a sex specific manner, building on animal studies where specific proteins have been found to play sex-specific roles in pain processing, and certain drugs have been found to have sex-specific effects in terms of analgesia (reviewed by (107)).While there are clearly also many similarities, it is important that clinicians begin to recognize these differences in clinical assessment and awareness of risks. While more research is required, these differences may also eventually inform choice of medical treatment. For example, if more MCP in women is associated with immune function, drugs that influence this may have specific long-term effects (positive or negative) in women and not in men. Opioids are known to adversely affect immune function (105,106). These differences may also inform the search for new, or re-purposed drugs in a sex specific manner, building on animal studies where specific proteins have been found to play sex-specific roles in pain processing, and certain drugs have been found to have sex-specific effects in terms of analgesia (reviewed by (107)).

#### Limitations

These analyses were carried out using UK Biobank, which was used in our previous sex-combined MCP GWAS (17). In comparison to our previous analysis, sample size in each individual sex-stratified GWAS was lower, which leads to somewhat reduced power. However, sample sizes are still larger than many sex-combined GWAS analyses of chronic pain phenotypes, and meta-analysis of the sex-stratified GWAS outputs resulted in an increase in power to find MCP-associated SNPs overall. True replication is difficult due to heterogeneity in chronic pain phenotyping and available sample sizes of potential independent cohorts, but in an independent subset of the UK Biobank, a PRS constructed from each sex-stratified GWAS output was found to be significantly associated with CWP, a related but distinct chronic pain phenotype of interest.

Although this work was focused on sex differences in the genetics of MCP we examined only autosomal variation. An important extension of this work would be to assess genetic associations with X chromosome loci, which is likely to provide an additional heritability contribution and give a fuller picture of sex differences in MCP at the genetic level. Inclusion of the X chromosome in GWAS analyses is associated with specific methodological and statistical issues including lower quality genotyping array coverage of the X chromosome compared to autosomes, differences in how imputation needs to be implemented, differences in X chromosome dosage between the sexes leading to differences in population genetics/demographic history of the X chromosome relative to the autosomes, X inactivation, and changes to quality control required (and differing QC protocols between sexes) (108,109). We aim to address this in future work as we adapt our BOLT-LMM pipeline and downstream analysis pathway.

## Conclusions

Sex differences in chronic pain likely have, at least in part, a genetic basis and the study of complex traits such as chronic pain is likely to benefit from “sex-aware” analytical approaches. This study comprises one of the largest sex-stratified genetic analyses of a chronic pain phenotype, and highlights sex-specific MCP-associated SNPs, genomic risk loci, genes, genetic correlations, and patterns of tissue expression.

Sex-stratified GWAS can also provide an increase in power if heterogeneity in effects of trait-associated variants is seen between the sexes. Here, 24 novel genes and 11 novel independent lead SNPs were associated with MCP, in addition to the findings from previous non sex-stratified work, further contributing to understanding of genetic variation in chronic pain.

Genetic correlation results indicate possible sex-specific pleiotropy, including differing genetic correlations between certain psychiatric disorders and traits and chronic pain in women compared to men, and between ASD and chronic pain in men compared to women. In contrast, genetic correlations between psychiatric disorders and traits including MDD and depressive symptoms remain similar between the sexes and to sex-combined analyses. This is also the first study to use novel GWAS output on studies of suicide, and of RDoC mental health traits such as mood instability and anhedonia in genetic correlation analyses with chronic pain and provide an important insight into shared genetic factors between these comorbidities of chronic pain and MCP.

Almost all sex-specific MCP-associated genes show expression in the DRG, with one male-specific locus *AMIGO3* found to be DRG-specific, indicating a potential specialised role in nociception. Many MCP-associated loci in both sexes have been linked to immune function.

Overall, findings indicate significant sex differences in chronic pain at multiple levels, from SNP-level to transcriptomic. Results support theories of strong nervous system and immune involvement in chronic pain in both sexes and can potentially inform development of novel treatment approaches in future, and further add to understanding of the physiology of chronic pain.

## Data Availability

UK Biobank data are available upon application to UK Biobank. GWAS summary statistic outputs will be made available through University of Glasgow Enlighten upon manuscript publication and upon request. Mouse transcriptomic data are freely accessible at mousebrain.org.

## References

1. Merskey H, Bogduk N. Classification of Chronic Pain. IASP Pain Terminology. 1994. 240 p.

2. IASP. International Association for the Study of Pain [Internet]. [cited 2020 Apr 14]. Available from: https://www.iasp-pain.org/

3. Nicholasa M, Vlaeyenb JWS, Riefe W, Barkee A, Azizf Q, Benolielg R, et al. The IASP classification of chronic pain for ICD-11: Chronic primary pain. Pain. 2019;160(1):53–9.

4. Gureje O, Von Korff M, Kola L, Demyttenaere K, He Y, Posada-Villa J, et al. The relation between multiple pains and mental disorders: Results from the World Mental Health Surveys. Pain. 2008;135(1–2):82–91.

5. Von Korff M, Crane P, Lane M, Miglioretti DL, Simon G, Saunders K, et al. Chronic spinal pain and physical-mental comorbidity in the United States: Results from the national comorbidity survey replication. Pain. 2005;113(3):331–9.

6. Santos-Eggimann B, Wietlisbach V, Rickenbach M, Paccaud F, Gutzwiller F. One-year prevalence of low back pain in two Swiss regions. Estimates from the population participating in the 1992-1993 MONICA project. Spine (Phila Pa 1976). 2000;25(19):2473–9.

7. Palmer KT, Walsh K, Bendall H, Cooper C, Coggon D. Back pain in Britain: Comparison of two prevalence surveys at an interval of 10 years. Br Med J. 2000;320(7249):1577–8.

8. Goldberg DS, McGee SJ. Pain as a global public health priority. BMC Public Health [Internet]. 2011;11(1):770. Available from: http://www.biomedcentral.com/1471-2458/11/770

9. Breivik H, Collett B, Ventafridda V, Cohen R, Gallacher D. Survey of chronic pain in Europe: Prevalence, impact on daily life, and treatment. Eur J Pain. 2006;10(4):287–333.

10. James SL, Abate D, Abate KH, Abay SM, Abbafati C, Abbasi N, et al. Global, regional, and national incidence, prevalence, and years lived with disability for 354 Diseases and Injuries for 195 countries and territories, 1990-2017: A systematic analysis for the Global Burden of Disease Study 2017. Lancet. 2018;1789–858.

11. Von Korff M, Ormel J, Keefe FJ, Dworkin SF. Grading the severity of chronic pain. Pain [Internet]. 1992;50(1092):133–49. Available from: http://www.ncbi.nlm.nih.gov/pubmed/1408309

12. Hocking LJ, Morris AD, Dominiczak AF, Porteous DJ, Smith BH. Heritability of chronic pain in 2195 extended families. Eur J Pain (United Kingdom). 2012;16(7):1053–63.

13. Junqueira DRG, Ferreira ML, Refshauge K, Maher CG, Hopper JL, Hancock M, et al. Heritability and lifestyle factors in chronic low back pain: Results of the Australian Twin Low Back Pain Study (The AUTBACK study). Eur J Pain (United Kingdom). 2014;18(10):1410–8.

14. Williams FMK, Spector TD, MacGregor AJ. Pain reporting at different body sites is explained by a single underlying genetic factor. Rheumatology. 2010;49(9):1753–5.

15. McIntosh AM, Hall LS, Zeng Y, Adams MJ, Gibson J, Wigmore E, et al. Genetic and Environmental Risk for Chronic Pain and the Contribution of Risk Variants for Major Depressive Disorder: A Family-Based Mixed-Model Analysis. PLoS Med. 2016;13(8):1–17.

16. Suri P, Palmer MR, Tsepilov YA, Freidin MB, Boer CG, Yau MS, et al. Genome-wide meta-analysis of 158,000 individuals of European ancestry identifies three loci associated with chronic back pain. 2018;48(7):1–23.

17. Johnston KJA, Adams MJ, Nicholl BI, Ward J, Strawbridge RJ, Ferguson A, et al. Genome-wide association study of multisite chronic pain in UK Biobank. PLoS Genet. 2019;15(6):1–22.

18. Rawlik K, Canela-Xandri O, Tenesa A. Evidence for sex-specific genetic architectures across a spectrum of human complex traits. Genome Biol [Internet]. 2016;17(1):1–8. Available from: http://dx.doi.org/10.1186/s13059-016-1025-x

19. Khramtsova EA, Davis LK, Stranger BE. The role of sex in the genomics of human complex traits. Nat Rev Genet [Internet]. 2019;20(3):173–90. Available from: http://dx.doi.org/10.1038/s41576-018-0083-1

20. Xu X, Coats JK, Yang CF, Wang A, Ahmed OM, Alvarado M, et al. Modular genetic control of sexually dimorphic behaviors. Cell [Internet]. 2012;148(3):596–607. Available from: http://dx.doi.org/10.1016/j.cell.2011.12.018

21. Quinn MA, Cidlowski JA. Endogenous hepatic glucocorticoid receptor signaling coordinates sex-biased inflammatory gene expression. FASEB J. 2016;30(2):971–82.

22. McCormick H, Young PE, Hur SSJ, Booher K, Chung H, Cropley JE, et al. Isogenic mice exhibit sexually-dimorphic DNA methylation patterns across multiple tissues. BMC Genomics. 2017;18(1):1–9.

23. Gilks WP, Abbott JK, Morrow EH. Sex differences in disease genetics: Evidence, evolution, and detection. Trends Genet [Internet]. 2014;30(10):453–63. Available from: http://dx.doi.org/10.1016/j.tig.2014.08.006

24. Ge T, Chen CY, Neale BM, Sabuncu MR, Smoller JW. Phenome-wide heritability analysis of the UK Biobank. PLoS Genet. 2017;13(4):1–21.

25. Rahmioglu N, MacGregor S, Drong AW, Hedman Å K, Harris HR, Randall JC, et al. Genome-wide enrichment analysis between endometriosis and obesity-related traits reveals novel susceptibility loci. Hum Mol Genet. 2015;24(4):1185–99.

26. Hall E, Volkov P, Dayeh T, Esguerra JL O. S Salö S, Eliasson L, et al. Sex differences in the genome-wide DNA methylation pattern and impact on gene expression, microRNA levels and insulin secretion in human pancreatic islets. Genome Biol. 2014;15(12):522.

27. Kukurba KR, Parsana P, Balliu B, Smith KS, Zappala Z, Knowles DA, et al. Impact of the X chromosome and sex on regulatory variation. Genome Res. 2016;26(6):768–77.

28. Yao C, Joehanes R, Johnson AD, Huan T, Esko T, Ying S, et al. Sex- and age-interacting eQTLs in human complex diseases. Hum Mol Genet [Internet]. 2013 Nov 15;23(7):1947–56. Available from: https://doi.org/10.1093/hmg/ddt582

29. Kósa JP, Balla B, Speer G, Kiss J, Borsy A, Podani J, et al. Effect of menopause on gene expression pattern in bone tissue of nonosteoporotic women. Menopause. 2009;16(2):367–77.

30. Gomez-Santos C, Hernandez-Morante JJ, Margareto J, Larrarte E, Formiguera X, Martínez CM, et al. Profile of adipose tissue gene expression in premenopausal and postmenopausal women: Site-specific differences. Menopause. 2011;18(6):675–84.

31. Mitra I, Tsang K, Ladd-Acosta C, Croen LA, Aldinger KA, Hendren RL, et al. Pleiotropic Mechanisms Indicated for Sex Differences in Autism. PLoS Genet. 2016;12(11):1–27.

32. Bartley EJ, Fillingim RB. Sex differences in pain: A brief review of clinical and experimental findings. Br J Anaesth. 2013;111(1):52–8.

33. Fillingim RB, King CD, Ribeiro-Dasilva MC, Rahim-Williams B, Riley JL. Sex, Gender, and Pain: A Review of Recent Clinical and Experimental Findings. J Pain [Internet]. 2009;10(5):447–85. Available from: http://dx.doi.org/10.1016/j.jpain.2008.12.001

34. Fillingim RB. Biopsychosocial contributions to sex differences in pain. BJOG An Int J Obstet Gynaecol. 2015;122(6):769.

35. El-Shormilisy N, Strong J, Meredith PJ. Associations among gender, coping patterns and functioning for individuals with chronic pain: A systematic review. Pain Res Manag. 2015;20(1):48–55.

36. Sorge RE, Mapplebeck JCS, Rosen S, Beggs S, Taves S, Alexander JK, et al. Different immune cells mediate mechanical pain hypersensitivity in male and female mice. Nat Neurosci. 2015;18(8):1081–3.

37. Sorge RE, Totsch SK. Review Sex Differences in Pain. 2017;1281(February 2016):1271–81.

38. Klein SL, Flanagan KL. Sex differences in immune responses. Nat Rev Immunol. 2016;16(10):626–38.

39. Smith BH, Campbell A, Linksted P, Fitzpatrick B, Jackson C, Kerr SM, et al. Cohort profile: Generation scotland: Scottish family health study (GS: SFHS). The study, its participants and their potential for genetic research on health and illness. Int J Epidemiol. 2013;

40. Macfarlane GJ, Barnish MS, Jones GT. Persons with chronic widespread pain experience excess mortality: longitudinal results from UK Biobank and meta-analysis. Ann Rheum Dis [Internet]. 2017;76(11):1815–22. Available from: http://ard.bmj.com/lookup/doi/10.1136/annrheumdis-2017-211476

41. Kamaleri Y, Natvig B, Ihlebaek CM, Benth JS, Bruusgaard D. Number of pain sites is associated with demographic, lifestyle, and health-related factors in the general population. Eur J Pain. 2008;12(6):742–8.

42. Bycroft C, Freeman C, Petkova D, Band G, Elliott LT, Sharp K, et al. The UK Biobank resource with deep phenotyping and genomic data. Nature. 2018;562(7726):203–9.

43. Loh PR, Tucker G, Bulik-Sullivan BK, Vilhjálmsson BJ, Finucane HK, Salem RM, et al. Efficient Bayesian mixed-model analysis increases association power in large cohorts. Nat Genet [Internet]. 2015;47(3):284–90. Available from: http://dx.doi.org/10.1038/ng.3190

44. Watanabe K, Taskesen E, Van Bochoven A, Posthuma D. Functional mapping and annotation of genetic associations with FUMA. Nat Commun [Internet]. 2017;8(1):1–10. Available from: http://dx.doi.org/10.1038/s41467-017-01261-5

45. de Leeuw CA, Mooij JM, Heskes T, Posthuma D. MAGMA: Generalized Gene-Set Analysis of GWAS Data. PLoS Comput Biol. 2015;11(4).

46. Aguet F, Brown AA, Castel SE, Davis JR, He Y, Jo B, et al. Genetic effects on gene expression across human tissues. Nature. 2017;550(7675):204–13.

47. Willer CJ, Li Y, Abecasis GR, Overall P. METAL?: fast and efficient meta-analysis of genomewide association scans. Bioinformatics. 2010;26(17):2190–1.

48. Ray P, Torck A, Quigley L, Wangzhou A, Neiman M, Rao C, et al. Comparative transcriptome profiling of the human and mouse dorsal root ganglia. Pain. 2018;159(7):1325–45.

49. Zeisel A, Hochgerner H, Lönnerberg P, Johnsson A, Memic F, van der Zwan J, et al. Molecular Architecture of the Mouse Nervous System. Cell. 2018;174(4):999-1014.e22.

50. Won H, Huang J, Opland CK, Hartl CL, Geschwind DH. Human evolved regulatory elements modulate genes involved in cortical expansion and neurodevelopmental disease susceptibility. Nat Commun [Internet]. 2019;10(1):1–11. Available from: http://dx.doi.org/10.1038/s41467-019-10248-3

51. Zheng J, Erzurumluoglu AM, Elsworth BL, Kemp JP, Howe L, Haycock PC, et al. LD Hub: A centralized database and web interface to perform LD score regression that maximizes the potential of summary level GWAS data for SNP heritability and genetic correlation analysis. Bioinformatics. 2017;33(2):272–9.

52. Bulik-Sullivan B, Loh PR, Finucane HK, Ripke S, Yang J, Patterson N, et al. LD score regression distinguishes confounding from polygenicity in genome-wide association studies. Nat Genet [Internet]. 2015;47(3):291–5. Available from: http://dx.doi.org/10.1038/ng.3211

53. Bulik-sullivan B, Finucane HK, Anttila V, Gusev A, Day FR, Case T, et al. An atlas of genetic correlations across human diseases and traits. Nat Publ Gr [Internet]. 2015;47(11):1236–41. Available from: http://dx.doi.org/10.1038/ng.3406

54. Dudbridge F. Power and Predictive Accuracy of Polygenic Risk Scores. PLoS Genet. 2013;9(3).

55. Watanabe K, Taskesen E, Van Bochoven A, Posthuma D. Functional mapping and annotation of genetic associations with FUMA. Nat Commun. 2017;8(1):1–10.

56. Winkler TW, Justice AE, Graff M, Barata L, Feitosa MF, Chu S, et al. The Influence of Age and Sex on Genetic Associations with Adult Body Size and Shape: A Large-Scale Genome-Wide Interaction Study. Vol. 11, PLoS Genetics. 2015. 1–42 p.

57. Randall JC, Winkler TW, Kutalik Z, Berndt SI, Jackson AU, Monda KL, et al. Sex-stratified Genome-wide Association Studies Including 270,000 Individuals Show Sexual Dimorphism in Genetic Loci for Anthropometric Traits. PLoS Genet. 2013;9(6).

58. Myers RA, Scott NM, Gauderman WJ, Qiu W, Mathias RA, Romieu I, et al. Genome-wide interaction studies reveal sex-specific asthma risk alleles. Hum Mol Genet. 2014;23(19):5251–9.

59. Prendergast L, van Vuuren C, Kaczmarczyk A, Doering V, Hellwig D, Quinn N, et al. Premitotic Assembly of Human CENPs -T and -W switches centromeric Chromatin to a mitotic state. PLoS Biol. 2011;9(6):1–12.

60. Lee S, Gang J, Jeon SB, Choo SH, Lee B, Kim YG, et al. Molecular cloning and functional analysis of a novel oncogene, cancer-upregulated gene 2 (CUG2). Biochem Biophys Res Commun. 2007;360(3):633–9.

61. Chun Y, Park B, Koh W, Lee S, Cheon Y, Kim R, et al. New centromeric component CENP-W Is an RNA-associated nuclear matrix protein that interacts with nucleophosmin/B23 protein. J Biol Chem. 2011;286(49):42758–69.

62. Schwarz M, Andrade-Navarro MA, Gross A. Mitochondrial carriers and pores: Key regulators of the mitochondrial apoptotic program? Apoptosis. 2007;12(5):869–76.

63. Willer CJ, Speliotes EK, Loos RJF, Li S, Lindgren CM, Heid IM, et al. Six new loci associated with body mass index highlight a neuronal influence on body weight regulation. Nat Genet. 2009;41(1):25–34.

64. Yu K, Ganesan K, Tan LK, Laban M, Wu J, Xiao DZ, et al. A precisely regulated gene expression cassette potently modulates metastasis and survival in multiple solid cancers. PLoS Genet. 2008;4(7).

65. Renström F, Payne F,Nordström A, Brito EC, Rolandsson O, Hallmans G, et al. Replication and extension of genome-wide association study results for obesity in 4923 adults from northern Sweden. Hum Mol Genet. 2009;18(8):1489–96.

66. Backofen B, Jacob R, Serth K, Gossler A, Naim HY, Leeb T. Cloning and characterization of the mammalian-specific nicolin 1 gene (NICN1) encoding a nuclear 24 kDA protein. Eur J Biochem. 2002;269(21):5240–5.

67. Alderson TRR, Kim JHH, Markley JLL. Dynamical Structures of Hsp70 and Hsp70-Hsp40 Complexes. Structure [Internet]. 2016;24(7):1014–30. Available from: http://dx.doi.org/10.1016/j.str.2016.05.011

68. Pencheva N, Tran H, Buss C, Huh D, Drobnjak M, Busam K, et al. Convergent multi-mRNA Targeting of ApoE Drives LRP1/LRP8-Dependent Melanoma Metastasis and Angiogenesis. Cell. 2012;151(5):1068–82.

69. Rauch U, Karthikeyan L, Maurel P, Margolis RU, Margolis RK. Cloning and primary structure of neurocan, a developmentally regulated, aggregating chondroitin sulfate proteoglycan of brain. J Biol Chem. 1992;267(27):19536–47.

70. Cichon S, Mühleisen TW, Degenhardt FA, Mattheisen M, Miró X, Strohmaier J, et al. Genome-wide association study identifies genetic variation in neurocan as a susceptibility factor for bipolar disorder. Am J Hum Genet. 2011;88(3):372–81.

71. Miró X, Meier S, Dreisow ML, Frank J, Strohmaier J, Breuer R, et al. Studies in humans and mice implicate neurocan in the etiology of mania. Am J Psychiatry. 2012;169(9):982–90.

72. Zhu CH, Kim J, Shay JW, Wright WE. SGNP: An essential stress granule/nucleolar protein potentially involved in 5.8s rRNA processing/transport. PLoS One. 2008;3(11).

73. Islam TC, Asplund AC, Lindvall JM, Nygren L, Liden J, Kimby E, et al. High level cannabinoid receptor 1, resistance of regulator G protein signaling 13 and differential expression of Cyclin D1 in mantle cell lymphoma. Leukemia. 2003;17(9):1880–90.

74. Nozawa T, Sano S, Minowa-Nozawa A, Toh H, Nakajima S, Murase K, et al. TBC1D9 regulates TBK1 activation through Ca2+ signaling in selective autophagy. Nat Commun [Internet]. 2020;11(1):1–16. Available from: http://dx.doi.org/10.1038/s41467-020-14533-4

75. Verploegen S, Lammers JWJ, Koenderman L, Coffer PJ. Identification and characterization of CKLiK, a novel granulocyte Ca++/calmodulin-dependent kinase. Blood. 2000;96(9):3215–23.

76. Bhattaram P, Penzo-Méndez A, Sock E, Colmenares C, Kaneko KJ, Vassilev A, et al. Organogenesis relies on SoxC transcription factors for the survival of neural and mesenchymal progenitors. Nat Commun. 2010;1(1):1–12.

77. Jay P, Gozé C, Marsollier C, Taviaux S, Hardelin JP, Koopman P, et al. The human SOX11 Gene: Cloning, chromosomal assignment and tissue expression. Genomics. 1995;29(2):541–5.

78. Haslinger A, Schwarz TJ, Covic M, Chichung Lie D. Expression of Sox11 in adult neurogenic niches suggests a stage-specific role in adult neurogenesis. Eur J Neurosci. 2009;29(11):2103–14.

79. Bergsland M, Werme M, Malewicz M, Perlmann T, Muhr J. The establishment of neuronal properties is controlled by Sox4 and Sox11. Genes Dev. 2006;20(24):3475–86.

80. Larson BL, Ylostalo J, Lee RH, Gregory C, Prockop DJ. Sox11 is expressed in early progenitor human multipotent stromal cells and decreases with extensive expansion of the cells. Tissue Eng - Part A. 2010;16(11):3385–94.

81. Tsurusaki Y, Koshimizu E, Ohashi H, Phadke S, Kou I, Shiina M, et al. De novo SOX11 mutations cause Coffin-Siris syndrome. Nat Commun. 2014;5:1–7.

82. Kuryshev VY, Vorobyov E, Zink D, Schmitz J, Rozhdestvensky TS, Münstermann E, et al. An anthropoid-specific segmental duplication on human chromosome 1q22. Genomics. 2006;88(2):143–51.

83. Kikuno R, Nagase T, Ishikawa K, Hirosawa M, Miyajima N, Tanaka A, et al. Prediction of the Coding Sequences of Unidentified Human Genes. XIV. The Complete Sequences of 100 New cDNA Clones from Brain Which Code for Large Proteins in vitro. DNA Res. 1999;205:197–205.

84. Bisogno T, Howell F, Williams G, Minassi A, Cascio MG, Ligresti A, et al. Cloning of the first sn1-DAG lipases points to the spatial and temporal regulation of endocannabinoid signaling in the brain. J Cell Biol. 2003;163(3):463–8.

85. Manitt C, Mimee A, Eng C, Pokinko M, Stroh T, Cooper HM, et al. The Netrin Receptor DCC Is Required in the Pubertal Organization of Mesocortical Dopamine Circuitry. J Neurosci. 2011;31(23):8381–94.

86. Arakawa H. Netrin-1 and its receptors in tumorigenesis. Nat Rev Cancer. 2004;4(12):978–87.

87. Srour M, Rivière JB, Pham JMT, Dubé MP, Girard S, Morin S, et al. Mutations in DCC cause congenital mirror movements. Science (80-). 2010;328(5978):592.

88. Savage JE, Jansen PR, Stringer S, Watanabe K, Bryois J, De Leeuw CA, et al. Genome-wide association meta-analysis in 269,867 individuals identifies new genetic and functional links to intelligence. Nat Genet. 2018;50(7):912–9.

89. Wright KM, Rand KA, Kermany A, Noto K, Curtis D, Garrigan D, et al. A prospective analysis of genetic variants associated with human lifespan. G3 Genes, Genomes, Genet. 2019;9(9):2863–78.

90. Strawbridge RJ, Ward J, Ferguson A, Graham N, Shaw RJ, Cullen B, et al. Identification of novel genome-wide associations for suicidality in UK Biobank, genetic correlation with psychiatric disorders and polygenic association with completed suicide. EBioMedicine [Internet]. 2019;41:517–25. Available from: https://doi.org/10.1016/j.ebiom.2019.02.005

91. Ward J, Tunbridge EM, Sandor C, Lyall LM, Ferguson A, Strawbridge RJ, et al. The genomic basis of mood instability: identification of 46 loci in 363,705 UK Biobank participants, genetic correlation with psychiatric disorders, and association with gene expression and function. Mol Psychiatry [Internet]. 2019; Available from: http://dx.doi.org/10.1038/s41380-019-0439-8

92. Okifuji A, Benham B. Suicidal and Self-Harm Behaviors in Chronic. J Appl Biobehav Res. 2011;16(2):57–77.

93. Nock MK, Borges G, Bromet EJ, Cha CB, Kessler RC, Lee S. Suicide and Suicidal Behavior Matthew. Epidemiol Rev. 2008;30(1):133–54.

94. Cheatle MD. Depression, chronic pain, and suicide by overdose: On the edge. Pain Med. 2011;12(SUPPL. 2):43–8.

95. Tang NKY, Crane C. Suicidality in chronic pain: A review of the prevalence, risk factors and psychological links. Psychol Med. 2006;36(5):575–86.

96. Triñanes Y, González-Villar A, Gómez-Perretta C, Carrillo-de-la-Peña MT. Suicidality in Chronic Pain: Predictors of Suicidal Ideation in Fibromyalgia. Pain Pract. 2015;15(4):323–32.

97. Smith MT, Edwards RR, Robinson RC, Dworkin RH. Suicidal ideation, plans, and attempts in chronic pain patients: Factors associated with increased risk. Pain. 2004;111(1–2):201–8.

98. Jung SJ, Winning A, Roberts AL, Nishimi K, Chen Q, Gilsanz P, et al. Posttraumatic stress disorder symptoms and television viewing patterns in the Nurses’ Health Study II: A longitudinal analysis. PLoS One. 2019;14(3):1–13.

99. Liberzon I, King AP, Ressler KJ, Almli LM, Zhang P, Ma ST, et al. Interaction of the ADRB2 gene polymorphism with childhood trauma in predicting adult symptoms of posttraumatic stress disorder. JAMA Psychiatry. 2014;71(10):1174–82.

100. Bierut LJ, Strickland JR, Thompson JR, Afful SE, Cottler LB. Drug use and dependence in cocaine dependent subjects, community-based individuals, and their siblings. Drug Alcohol Depend. 2008;95(1–2):14–22.

101. Luo Z, Alvarado GF, Hatsukami DK, Johnson EO, Bierut LJ, Breslau N. Race differences in nicotine dependence in the Collaborative Genetic study of Nicotine Dependence (COGEND). Nicotine Tob Res. 2008;10(7):1223–30.

102. Baker DG, Nash WP, Litz BT, Geyer MA, Risbrough VB, Nievergelt CM, et al. Predictors of Risk and Resilience for Posttraumatic Stress Disorder Among Ground Combat Marines: Methods of the Marine Resiliency Study. Prev Chronic Dis. 2012;9(5):1–11.

103. Kimbrel NA, Hauser MA, Garrett M, Ashley-Koch A, Liu Y, Dennis MF, et al. Effect of the APOE ε4 allele and combat exposure on PTSD among Iraq/Afghanistan-era veterans. Depress Anxiety. 2015;32(5):307–15.

104. Nievergelt CM, Maihofer AX, Klengel T, Atkinson EG, Chen C-Y, …, et al. Largest genome-wide association study for PTSD identifies genetic risk loci in European and African ancestries and implicates novel biological pathways. bioRxiv [Internet]. 2018;(Nov). Available from: http://dx.doi.org/10.1101/458562

105. Liang X, Liu R, Chen C, Ji F, Li T. Opioid System Modulates the Immune Function: A Review Xuan. Transl Perioper Pain Med. 2016;1(1):5–13.

106. Diasso PDK, Birke H, Nielsen SD, Main KM, Højsted J, Sjøgren P, et al. The effects of long-term opioid treatment on the immune system in chronic non-cancer pain patients: A systematic review. Eur J Pain (United Kingdom). 2020;24(3):481–96.

107. Mogil JS. Qualitative sex differences in pain processing: emerging evidence of a biased literature. Nat Rev Neurosci 2020 [Internet]. 2020;1–13. Available from: https://www.nature.com/articles/s41583-020-0310-6 https://www.nature.com/articles/s41583-020-0310-6

108. König IR, Loley C, Erdmann J, Ziegler A. How to Include Chromosome X in Your Genome-Wide Association Study. Genet Epidemiol. 2014;

109. Accounting for sex in the genome. Nat Med. 2017;23(11):1243.

